# Global determinants of vector-targeted insecticide use in public health: a modeling and mapping analysis

**DOI:** 10.64898/2026.04.08.26350404

**Authors:** Patrick M. Heffernan, Henk van den Berg, Rajpal S. Yadav, Courtney C. Murdock, Jason R. Rohr

## Abstract

**Background:** Insecticides are the cornerstone of control for vectors of human pathogens, yet global patterns of deployment and their socioeconomic and environmental drivers are poorly characterized. Understanding where and why insecticides are used is essential for optimizing control efforts.

**Methods:** We analyzed annual national vector-targeted insecticide-use data spanning 123 countries (1990-2019) for insecticide-treated nets (ITNs), residual spraying (RS), space spraying (SS), and larviciding (LA). Generalized linear mixed models and hurdle models quantified associations between deployment and mosquito-borne disease incidence, human development index (HDI), population, temperature, and precipitation. Models were evaluated using repeated cross-validation and applied to generate downscaled predictions of insecticide use at subnational administrative regions globally.

**Findings:** Insecticide deployment increased with mosquito-borne disease incidence, but this response was stronger in higher-HDI countries, indicating that deployment depends on socioeconomic capacity as well as disease burden. Intervention types each exhibited distinct patterns. ITN and RS tracked *Anopheles-*borne disease incidence, whereas SS and LA tracked *Aedes*-borne disease incidence. ITN use increased with precipitation and temperature, whereas SS use decreased with precipitation. Mapping uncovered substantial within-country heterogeneity, highlighting regions where predicted deployment remains low relative to disease risk.

**Interpretation:** Insecticide deployment reflects epidemiological need and economic capacity, creating mismatches between risk and control. High-resolution mapping can support more equitable intervention allocation, guide insecticide resistance stewardship, and improve strategic planning as climate and urbanization reshape vector-borne disease risk.

**Funding:** JRR acknowledges support from NSF (DEB-2109293, ITE-2333795, BCS-2307944, CISE-2435758), Notre Dame Poverty Initiative, Berthiaume Institute for Precision Health, IL-IN Sea Grant, Frontier Research Foundation.

**Research in Context:** *Evidence before this study:* Insecticides are one of the main pillars for controlling arthropod vectors of human diseases, such as mosquitoes, but data on global deployment remain limited. We conducted a search of Google Scholar from 2000 through 2025 using the search term ‘global determinants insecticide use’ to identify global data on vector-targeted insecticide use. We included studies and datasets that quantified global use of vector-targeted insecticide interventions at a national scale and excluded studies restricted to individual locations or regions, single interventions, or non-vector targeted insecticide use. Two survey-based datasets (published in 2012 and 2021) contain information on national-level, self-reported, vector-targeted insecticide deployment for major intervention types including insecticide-treated nets, residual spraying, space spraying, and larviciding. The overall quality of these sources is moderate as not all countries were surveyed and not all countries reported data for every year. Additionally, the Net Mapping Project provides data on national-level insecticide-treated net procurement based on shipment and delivery data, rather than nation-reported retrospective data. Previous assessments of broad-scale trends in these datasets have revealed substantial variation in usage across global regions and among intervention types. For example, in the 2010s, insecticide use on the continent of Africa was dominated by an increasing proportion of insecticide-treated nets, whereas use in the Asia-Pacific, Latin America, and Caribbean regions was dominated by residual spraying, space spraying, and larviciding. Additionally, insecticide efficacy and coverage are known to be impacted by socioeconomic and environmental conditions. However, to our knowledge, no study has assessed national-level, vector-targeted insecticide deployment across key socioeconomic and environmental predictors.

*Added value of this study:* To our knowledge, this is the first global analysis and down-scaled mapping study of major vector-targeted interventions. By analyzing vector-targeted insecticide deployment datasets in response to national-level socioeconomic, environmental and mosquito-borne disease incidence predictors across 123 countries, we identify key deployment patterns in overall insecticide use as well as specific use in insecticide-treated nets, residual spraying, space spraying, and larviciding. We find that higher-HDI countries had the highest deployment, indicating the importance of socioeconomic capacity. We also find that deployment increased with population density and varied predictably with temperature and precipitation. Furthermore, by analyzing each insecticide intervention type separately, this study determines that each intervention type can exhibit distinct patterns of deployment. For example, space spraying is positively associated with *Aedes-*borne disease incidence, whereas insecticide-treated nets are positively associated with *Anopheles-*borne disease incidence. This study also provides the first global map of vector-targeted insecticide deployment at the administrative unit 2 level. The mapping uncovers substantial within-country heterogeneity for insecticide usage masked at the national scale.

*Implications of all the available evidence:* Strengthening global vector control, particularly in the face of growing insecticide resistance and shifting vector ranges, is of highest priority in public health. Our findings highlight that vector-targeted insecticide use depends on socioeconomic status in addition to environmental and epidemiological conditions, which may drive successful vector-control. The patterns uncovered here highlight should be incorporated into predictive spatial frameworks to anticipate current and future intervention needs. The downscaled global maps can be used to identify hotspots of insecticide demand, guide proactive resource allocation, and support insecticide resistance management before mismatches between disease risk and control capacity become more pronounced.

## Introduction

The impact of mosquito-borne diseases is immense, with over 50 million disability-adjusted life years (DALYs) lost to diseases such as malaria and dengue fever in 2017 alone.^1^ Malaria accounted for most of this burden (∼45 million DALYs), followed by dengue (∼2·9 million), lymphatic filariasis (∼1·4 million), yellow fever (∼0·3 million), and Zika (∼2·2 thousand). This burden is likely underestimated as these estimates do not include several emerging mosquito-borne diseases that have expanded in geographic range and public health importance in recent decades, such as West Nile virus and chikungunya. Further, tens of billions of United States dollars (USD) are spent annually on treatment and vector control,^2^ and this burden is projected to increase up to 14-fold as urban-adapted mosquitoes expand their historical geographical ranges.^2,3^ Thus, the global economic burden of these diseases is large and growing, with insecticides constituting the backbone of vector-control efforts for over a century.^4^

Primary insecticidal approaches for protecting people from vector-borne disease include the following: 1) Insecticide-treated nets (ITNs), which predominantly target adult *Anopheles* mosquitoes as well as sandflies; 2) Residual spraying (RS), which applies insecticides to indoor and outdoor surfaces to target resting mosquitoes, sandflies, and triatomes; 3) Space spraying (SS), which uses fogging to rapidly kill adult *Aedes* mosquitoes in flight during arbovirus outbreaks; and 4) Larviciding (LA), which targets immature mosquito life stages in aquatic habitats. Despite extensive investment, these tools face growing challenges that include poor coordination across methods and widespread insecticide resistance.^4^

To strengthen global vector control, the WHO launched the *Global Vector Control Response 2017–2030 (GVCR)*,^5^ which calls for integrated and evidence-based strategies. Critical to this effort is the ability to monitor and assess where and how insecticides are deployed, yet comprehensive global data have been limited. Newly compiled national-level datasets on annual insecticide use now provide an opportunity to evaluate global patterns and identify key drivers of deployment.^6,7^ Without such information, programs risk inefficient allocation, insufficient coverage in high-burden settings, and accelerated resistance.

Because insecticides are commercial and operational interventions rather than purely biological responses, their deployment is shaped not only by disease burden but also by social, economic, and environmental constraints and global donor support, especially for malaria control. Vector–borne diseases disproportionately affect low- and middle-income countries,^8^ yet large-scale insecticide programs, particularly RS and SS, require substantial infrastructure, logistics, and sustained financing. As a result, regions with the highest need may not always achieve the highest coverage, creating mismatches between disease risk and control capacity. Additionally, for *Aedes*-borne diseases such as dengue, insecticides are only one component of vector control. Many programs prioritize habitat source reduction and community-based interventions targeting aquatic sites that support juvenile mosquito development. Consequently, insecticide use represents only one dimension of *Aedes-*borne disease control efforts. Ecological conditions further modify insecticide use as temperature, humidity, and precipitation influence habitat suitability, mosquito abundance, and mosquito population dynamics.^9^ Environmental variation, such as seasonal duration and environmental suitability for transmission, can also alter the timing and coverage required for interventions.^10^ Together, these considerations suggest that insecticide deployment should vary systematically across development, population density, disease burden, and climate, and that these drivers may differ among intervention types.

Here, our primary goals are to identify the socioeconomic and climatic drivers of insecticide deployment targeting vectors and generate high-resolution maps to support targeting, planning, and evaluation of vector-control strategies. We also examine how these drivers relate to the deployment of each of the following major insecticide categories: ITN, SS, RS, and LA. Using national-level insecticide-use data,^6,7^ we identify the magnitude and direction of key predictors and their interactions and then generate predicted downscaled insecticide-use maps at the administrative region 2 (ADM2) scale (counties, districts, regions; as characterized by the Global Administrative Unit Layers by the Food and Agriculture Organization (FAO GAUL)^11^). As global datasets on demographic, socioeconomic, and climatic factors become increasingly detailed, there is growing value in producing finer-resolution maps of insecticide use. Downscaled maps can assist national programs and international agencies in understanding where control efforts are concentrated and how environmental and human factors shape the use of specific interventions. In particular, this national-level analysis may inform national programs that then go on to inform state-level and fine-scale municipality control. While there is environmental and socio-economic heterogeneity at fine-spatial scales not captured by a national-level analysis, these analyses are a sensible starting point based on the availability of national-level data.

## Methods

### Insecticide use dataset

We analyzed national-level insecticide-use data compiled by van den Berg et al. (2012, 2021)^6,7^ encompassing 123 countries from 1990 to 2019. The dataset reports annual national-level insecticide deployment for four major vector-control interventions: ITNs, RS, SS, and LA. To facilitate comparisons among intervention types, all insecticide use was expressed as standard spray coverage (SSC; km^2^ treated with active ingredient). Observations from years or countries without reported insecticide-use data were excluded. Because most reported insecticide use targeted malaria and dengue vectors, analyses focused on associations with *Anopheles-* and *Aedes-* borne disease incidence. Factory-treated ITNs were incorporated following the approach of van den Berg et al. (2021)^6^ using annual national-level delivery data from the Net Mapping Project.^12^

### Predictor variables

We evaluated associations between annual insecticide use and six predictors representing epidemiological, socioeconomic, demographic, and environmental factors: *Anopheles*-borne diseased incidence, *Aedes-*borne disease incidence, Human Development Index (HDI), human population density, mean monthly temperature, and mean monthly precipitation. Predictor values were matched to the corresponding year of each insecticide-use observation. Additionally, for the total annual insecticide use model, we tested mean monthly vapor pressure deficit in place of mean monthly temperature as a composite variable incorporating temperature and relative humidity.

Malaria incidence was used as a proxy for *Anopheles*-borne disease incidence, and dengue incidence was used as proxy for *Aedes-*borne disease incidence because these diseases dominate global insecticide-based control programs targeting their respective mosquito vectors. Annual disease incidence estimates were obtained from the Global Burden of Disease (GBD) database,^13^ HDI values were downloaded from the United Nations Development Programme,^14^ human population data were obtained from World Bank^15^ and converted to population density using country land area from the Central Intelligence Agency’s World Factbook,^16^ and climatic variables were derived from the European Centre for Medium-Range Weather Forecasts (ECMWF) ERA5-Land monthly aggregated dataset^17^ using Google Earth Engine.^18^

### Model development

We first modeled total annual insecticide use across all interventions (i.e., combining ITN, RS, SS, and LA) using a generalized linear mixed-effects model (GLMM) in the *glmmTMB*^19,20^ package in *R,*^21^ with country included as a random intercept and national population as an offset. Fixed effects included *Anopheles-*borne disease incidence, *Aedes*-borne disease incidence, HDI, population density, temperature, precipitation, and their interactions. Predictor selection was performed using *glmmLasso,*^22^ after which the final model was refit in *glmmTMB.* Model performance was evaluated using repeated cross-validation to assess predictive accuracy. Separate hurdle models were then developed for each intervention type, modeling both the probability of deployment and the magnitude of deployment among countries reporting use. Additional methodological details including predictor transformations, model selection, diagnostics, and cross-validation procedures, are provided in the *Supplementary Methods.* Marginal prediction plots for significant terms from the most parsimonious model were generated using the *sjPlot*^23^ and *ggplot*^24^ packages.

### Spatial prediction and mapping

To generate high-resolution global prediction maps, spatial datasets corresponding to each predictor were assembled at the administrative unit 2 (ADM2) level.^11^ Malaria incidence was obtained from the Malaria Atlas Project,^25^ dengue incidence was estimated from global force-of-infection maps^26^ with LandScan population data,^27^ and temperature and precipitation from the ERA5-Land dataset.^17^ Predicted insecticide use at the ADM2 level was generated from the final overall model using the *ggeffects* package^28^ coupled with spatial predictor datasets (see *Supplementary Methods* for details).

## Results

### Overall insecticide use model

A total of 1,658 country-year insecticide use observations from 123 countries (1990-2019) were included in the final overall model. A summary of the most salient associations is presented in Table 1 (see Table S1, Fig. 1, and Fig. S1 for more complete results).

**Figure 1.**
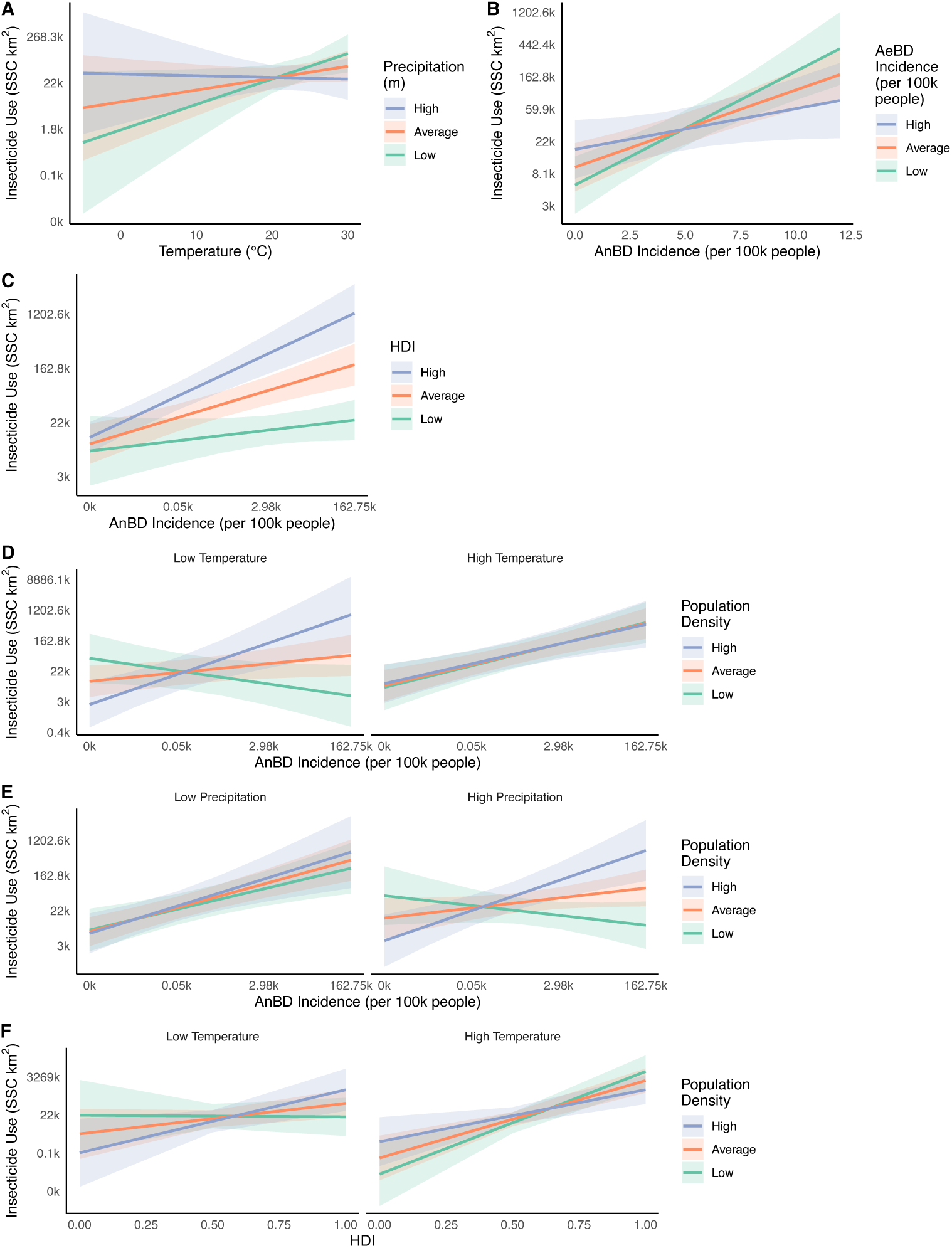
Significant three-way and two-way interactions from the overall annual insecticide use model, with precipitation (mean monthly, m; **A, E**), temperature (mean monthly, ℃; **A, D, F**), *Anopheles*-borne disease (AnBD) incidence (new cases divided by mid-year population; **B-E**), HDI (human development index; **C, F**), *Aedes*-borne disease (AeBD) incidence (new cases divided by mid-year population; **B**), and population density (**D-F**) as predictors. Insecticide use (SSC = standard spray coverage in km^2^), AnBD and AeBD incidences, and precipitation are all presented on natural log scales. Shown are best fit lines and 95% confidence bands. *High* and *Low* represent the +/- 1 standard deviation from the mean.

**Table 1 -.** Direction of mosquito-targeting insecticide use in overall model.

| Predictor | Direction of Overall Use |
| --- | --- |
| *Anopheles*-borne Disease | +<sup>a</sup> |
| *Aedes*-borne Disease | +/-<sup>b</sup> |
| HDI | + |
| Population Density | +/-<sup>c</sup> |
| Warm, Dry | + |
| Cool, Wet | + |
| *Note: These results indicate the most salient patterns. '+' indicates an increase in use; '-' indicates a decrease in use. For more detailed and nuanced patterns, refer to predictor tables.* |  |
| <sup>a</sup> Positive with high temperature |  |
| <sup>b</sup> Positive when *Anopheles*-borne disease incidence is low, negative when high |  |
| <sup>c</sup> Positive when *Anopheles*-borne disease incidence is high, negative when low |  |

Overall insecticide use was significantly associated with *Anopheles-* and *Aedes-*borne disease incidences, HDI, population density, temperature, and precipitation (Table S1). Insecticide use increased with temperature and tended to be higher in wetter countries (Fig. 1A). Insecticide use tended to increase with *Anopheles-*borne disease incidence and increase with *Aedes-*borne disease incidence in countries without *Anopheles*-borne disease (Fig. 1B). Among warmer countries, the relationship between insecticide use and *Anopheles*-borne disease incidence remained positive regardless of population density (Fig. 1D). Among cooler countries, however, this relationship was strongest where population density was high (Fig. 1D). The relationship between disease burden and insecticide deployment was strongly modified by HDI, with higher-HDI countries exhibiting greater increases in use with increasing incidence (Fig. 1C). Overall insecticide usage tended to increase with HDI, particularly in warmer and more densely populated countries (Fig. 1F). As these analyses do not incorporate temporal lags, associations between insecticide use and disease incidence should not be interpreted as evidence of intervention effectiveness. When replacing temperature with vapor pressure deficit, the results were similar for the main model (Table S6).

### Intervention-specific patterns

The intervention-specific hurdle models included 1,658 observations for each binomial model. Gaussian models included 803 observations for ITNs (90 countries), 1,268 for RS (103 countries), 754 for SS (72 countries), and 940 for LA (85 countries). Across intervention types, factors associated with the likelihood of use frequently differed from those associated with the magnitude of deployment, indicating distinct determinants of adoption versus scale (Table 2; Tables S2-S5).

**Table 2 -.** Direction of mosquito-targeting insecticide use in insecticide type hurdle models.

|  |  | ITN |  | RS |  | SS |  | LA |  |
| --- | --- | --- | --- | --- | --- | --- | --- | --- | --- |
| Predictor |  | Binomial | Gaussian | Binomial | Gaussian | Binomial | Gaussian | Binomial | Gaussian |
| *Anopheles*-borne Disease |  | +<sup>a</sup> | + | +<sup>b</sup> | +<sup>b</sup> | - |  | - | - |
| *Aedes*-borne Disease |  |  |  |  |  | + | + |  | + |
| HDI |  | + | + | +<sup>c</sup> | +<sup>c</sup> | + |  | + | + |
| Population Density |  | + |  | - | +<sup>e</sup> |  |  |  |  |
| Temperature |  | + | + | -<sup>d</sup> |  |  |  |  |  |
| Precipitation |  | + | + | - |  |  | - |  |  |
| ITN Use |  |  |  |  | +<sup>d</sup> |  |  |  |  |
| RS Use |  |  | -<sup>a</sup> |  |  |  |  |  | +<sup>b</sup> |
| SS Use |  |  |  |  |  |  |  | +<sup>g</sup> | +<sup>g</sup> |
| LA Use |  |  |  |  |  | + | +<sup>f</sup> |  |  |
| *Note: These results indicate the most salient patterns discussed in the main text. '+' indicates an increase in use; '-' indicates a decrease in use. A blank cell indicates no salient pattern in the model. For more detailed and nuanced patterns, refer to figures and predictor tables.* |  |  |  |  |  |  |  |  |  |
| <sup>a</sup> With high temperature |  |  |  |  |  |  |  |  |  |
| <sup>b</sup> With high human development index (HDI) |  |  |  |  |  |  |  |  |  |
| <sup>c</sup> With high *Anopheles*-borne disease incidence |  |  |  |  |  |  |  |  |  |
| <sup>d</sup> With high population density |  |  |  |  |  |  |  |  |  |
| <sup>e</sup> With high insecticide-treated net (ITN) use |  |  |  |  |  |  |  |  |  |
| <sup>f</sup> With low precipitation |  |  |  |  |  |  |  |  |  |
| <sup>g</sup> With low *Anopheles*-borne disease incidence |  |  |  |  |  |  |  |  |  |

The likelihood of ITN deployment increased with HDI, population density, temperature, precipitation, and *Anopheles-* borne disease incidence (when temperature was high), whereas deployment magnitude increased with HDI, temperature, precipitation, and *Anopheles-*borne disease incidence (Fig. 2). Similarly, both the likelihood and magnitude of RS deployment increased with *Anopheles-*borne disease incidence in higher-HDI countries (Fig. 3).

**Figure 2.**
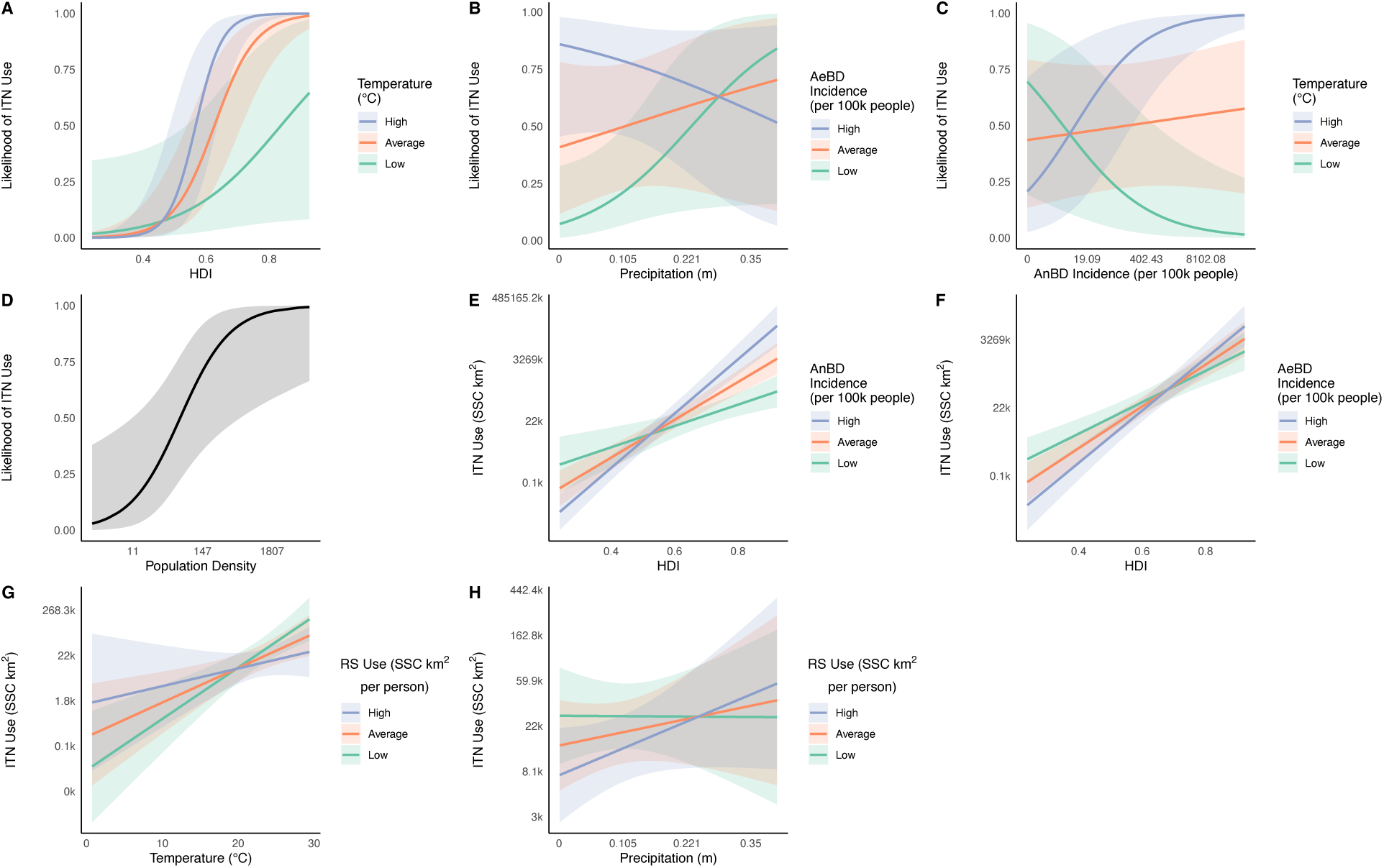
Marginal predictive plots displaying salient associations from the likelihood (**A-D**) and magnitude (**E-H**) components of the annual insecticide-treated net (ITN; SSC = standard spray coverage in km^2^) use hurdle model, with human development index (HDI; **A, E-F**), temperature (mean monthly, ℃; **A, C, G**), *Aedes*-borne disease incidence (AeBD; new cases divided by mid-year population; **B, F**), precipitation (mean monthly, m; **B, H**), *Anopheles*-borne disease incidence (AnBD; new cases divided by mid-year population; **C, E**), population density (people per km^2^; **D**), and residual spraying (RS) use (SSC per person; **G-H**) as predictors. ITN use, precipitation, AeBD, AnBD, population density, and RS use are presented on a natural log scale. The best fit lines and 95% confidence bands are displayed. In figures with interactions, *High* and *Low* represent the +/- 1 standard deviation from the mean.

**Figure 3.**
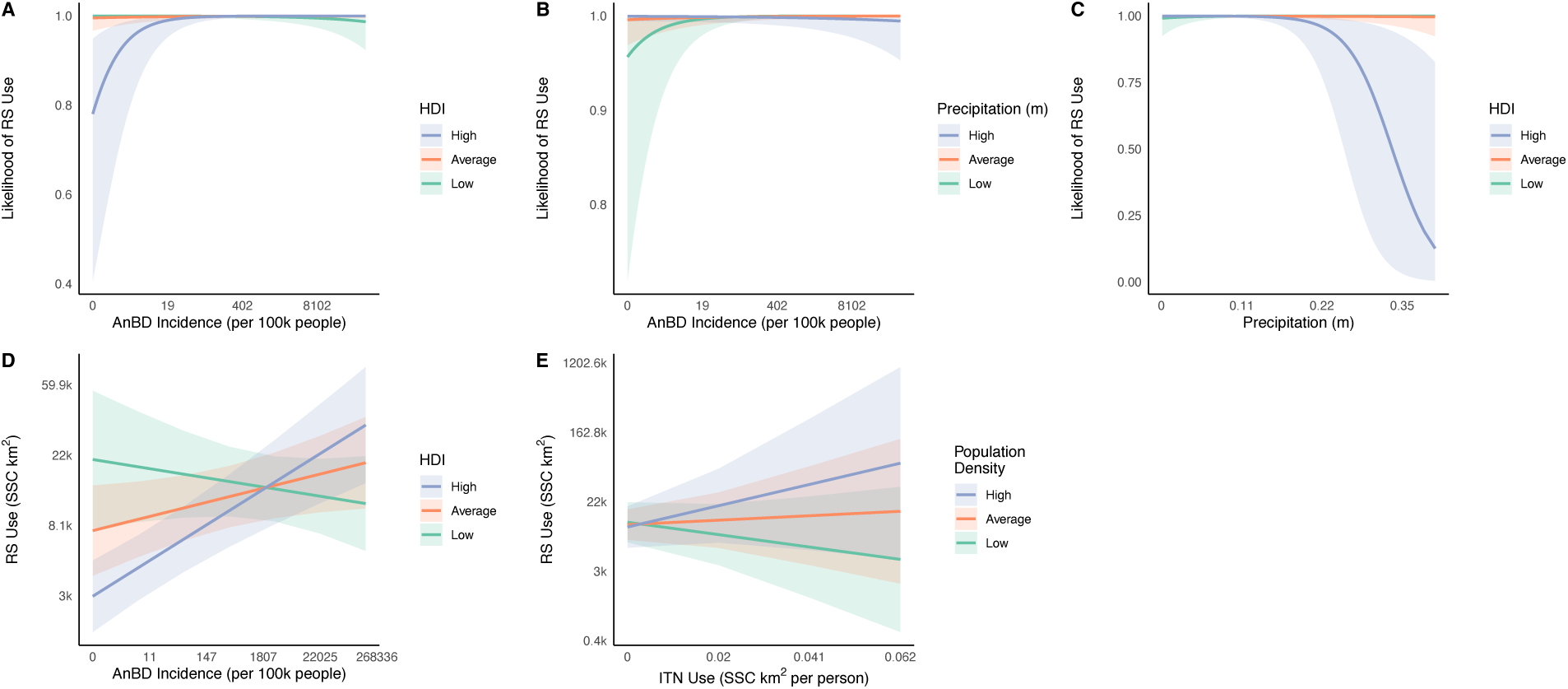
Marginal predictive plots displaying salient associations from the likelihood (**A-C**) and magnitude (**D-E**) components of the annual residual spraying (RS; SSC = standard spray coverage in km^2^) use hurdle model, with *Anopheles*-borne disease incidence (AnBD; new cases divided by mid-year population; **A-B, D**), human development index (HDI; **A, C-D**), precipitation (mean monthly, m; **B-C**), population density (people per km^2^; **E**), and insecticide-treated net (ITN) use (SSC per person; **E**) as predictors. RS use, ITN use, AnBD, precipitation, and population density are presented on a natural log scale. The best fit lines and 95% confidence bands are displayed. In figures with interactions, *High* and *Low* represent the +/- 1 standard deviation from the mean.

SS and LA exhibited patterns distinct from ITNs and RS. The likelihood of both interventions increased with HDI and decreased with *Anopheles-*borne disease incidence (Figs. 4B-C, 5B-D). SS and LA were the intervention types most likely to co-occur across settings (Fig. 4D, 5A), and their deployment magnitudes were positively correlated in countries with lower precipitation and *Anopheles-*borne disease incidence (Figs. 4G, 5G).

**Figure 4.**
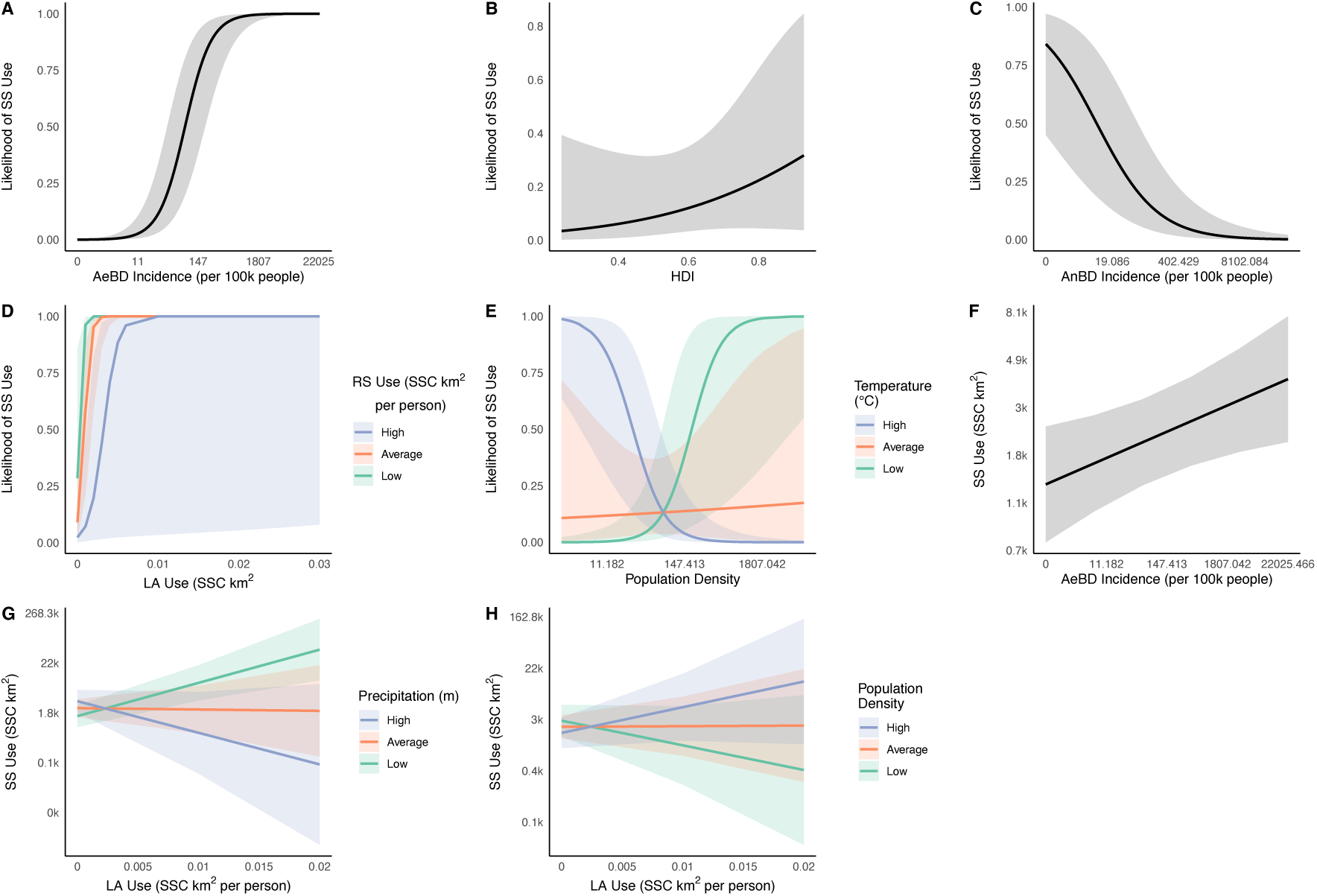
Marginal predictive plots displaying salient associations from the likelihood (**A-E**) and magnitude (**F-H**) components of the annual space spraying (SS; SSC = standard spray coverage in km^2^) use hurdle model, with *Aedes*-borne disease incidence (AeBD; new cases divided by mid-year population; **A, F**), human development index (HDI; **B**), *Anopheles*-borne disease incidence (AnBD; new cases divided by mid-year population; **C**), larvicide (LA) use (SSC per person; **D, G-H**), residual spraying (RS) use (SSC per person; **D**), population density (people per km^2^; **E, H**), temperature (mean monthly, ℃; **E**), and precipitation (mean monthly; m; **G**) as predictors. SS use, LA use, RS use, AeBD, AnBD, population density, and precipitation are presented on a natural log scale. The best fit lines and 95% confidence bands are displayed. In figures with interactions, *High* and *Low* represent the +/- 1 standard deviation from the mean.

**Figure 5.**
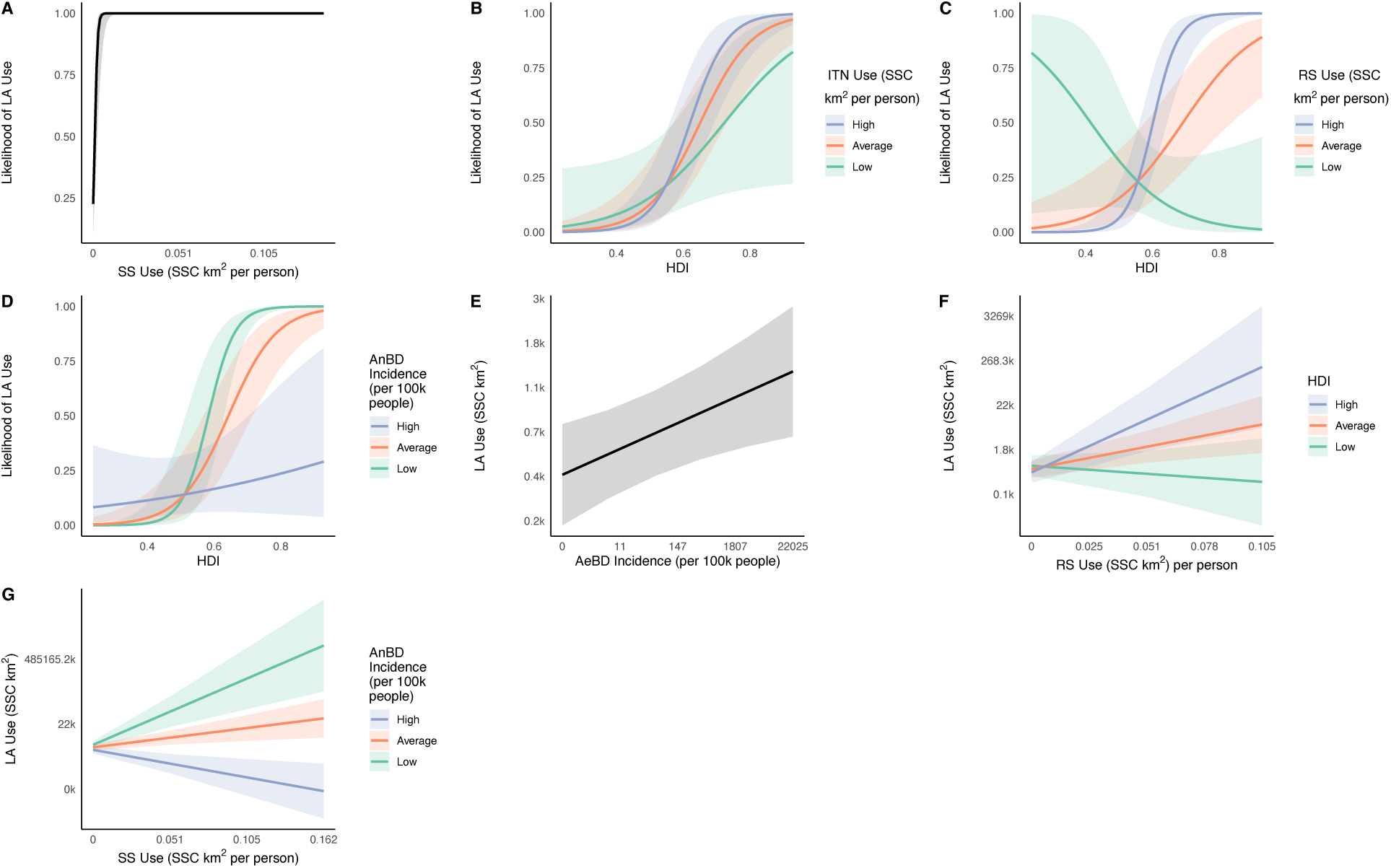
Marginal predictive plots displaying salient associations from the likelihood (**A-D**) and magnitude (**E-G**) components of the annual larvicide (LA; SSC = standard spray coverage in km^2^) use hurdle model, with space spraying (SS) use (SSC per person; **A, G**), human development index (HDI; **B-D, F**), insecticide-treated net (ITN) use (SSC per person; **B**), residual spraying (RS) use (SSC per person; **C**), *Anopheles*-borne disease incidence (AnBD; new cases divided by mid-year population; **D, G**), and *Aedes*-borne disease incidence (AeBD; new cases divided by mid-year population; **E**) as predictors. LA use, SS use, ITN use, RS use, AnBD, and AeBD are presented on a natural log scale. The best fit lines and 95% confidence bands are displayed. In figures with interactions, *High* and *Low* represent the +/- 1 standard deviation from the mean.

The likelihood and magnitude of SS deployment increased with *Aedes-*borne disease incidence (Fig. 4A, 4F), and the magnitude of LA deployment increased with *Aedes-*borne disease incidence (Fig. 5E).

Population density increased the likelihood of ITN and SS deployment (Figs. 2D, S4C), whereas RS deployment magnitude increased with population density only in countries with high ITN use (Fig. 3E). Temperature increased the likelihood and magnitude of ITN deployment (Figs. 2A, 2C, 2G) but decreased the likelihood of SS deployment in densely populated areas (Fig. 4E). Precipitation increased the likelihood and magnitude of ITN deployment (Fig. 2B, 2H) but was negatively associated with the likelihood or magnitude of RS and SS deployment (Figs. 3B, 4G).

### Spatial predictions

Predicted insecticide deployment was highest in densely populated metropolitan regions (e.g., Karachi and Delhi) and in regions with high *Anopheles*-borne disease burden despite relatively low socioeconomic development (e.g., parts of Burkina Faso and Zambia) (Fig. 6). Within countries, substantial spatial heterogeneity was evident, with deployment varying markedly among neighboring administrative units. Regions with high *Anopheles*-borne disease incidence generally exhibited greater predicted deployment than regions dominated by *Aedes*-borne disease, although areas with high burdens of both diseases also showed elevated predicted insecticide use. Averaging predictions to ADM0 and ADM1 occasionally obscured or even reversed local hotspots evident at the ADM2 level (e.g., Brazil and the United States; Fig. S5-S6). The downscaled maps identified subnational regions where predicted insecticide deployment remained low relative to disease burden, particularly across parts of sub-Saharan Africa and South Asia.

**Figure 6.**
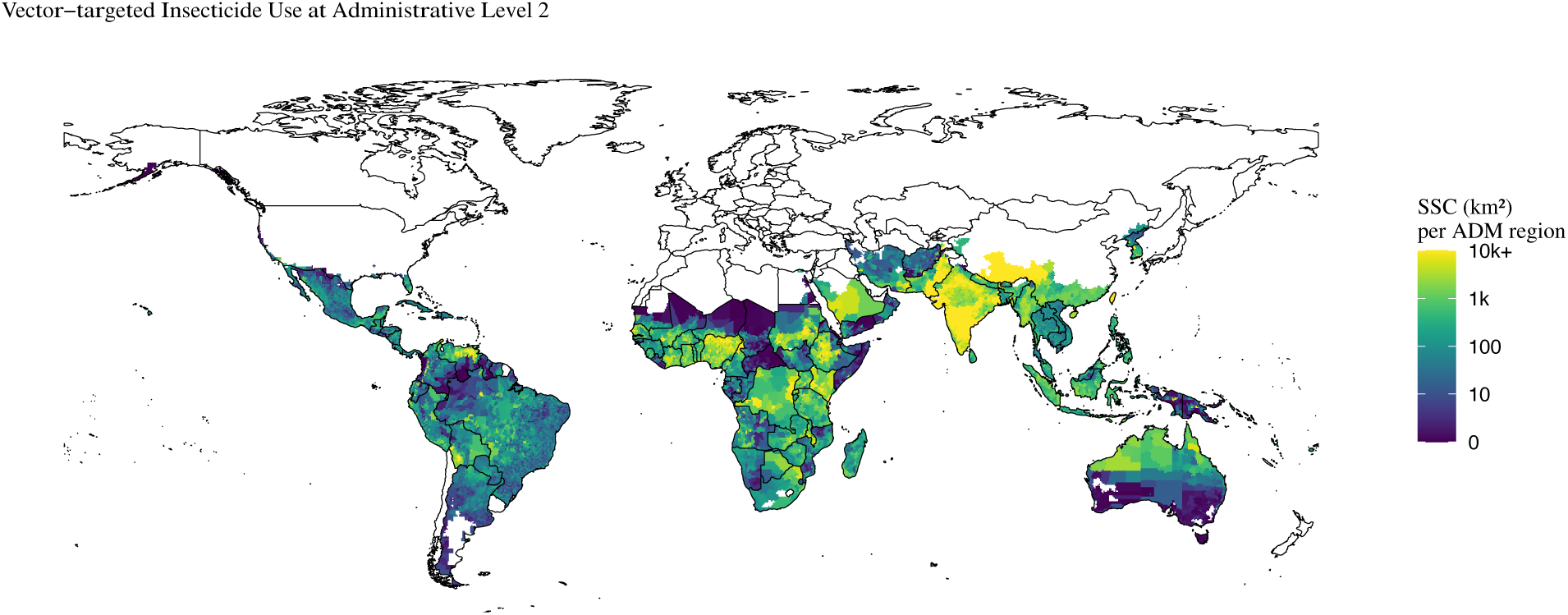
Predicted magnitude of total annual insecticide use (standard spray coverage in km^2^; including insecticide-treated nets, residual spraying, space spraying, and larviciding) for *Anopheles-* and *Aedes-*borne disease control at the administrative level 2 (ADM2).

## Discussion

This study provides, to our knowledge, the first global assessment of the socioeconomic, epidemiological, and environmental determinants of vector-targeted insecticide deployment. Three broad patterns emerged. First, insecticide deployment reflected not only mosquito-borne disease burden but also socioeconomic capacity, with higher-HDI countries scaling deployment more strongly in response to increasing disease incidence. Second, the four major intervention types exhibited distinct epidemiological and socioeconomic signatures, reflecting differences in their target vectors, transmission environments (urban vs. rural), and operational requirements. Third, downscaled predictions revealed substantial within-country heterogeneity that was obscured by national averages, demonstrating the value of high-resolution prediction for targeting vector-control resources. Together, these findings demonstrate that insecticide deployment reflects not only epidemiological need but also demographic, socioeconomic, and environmental conditions, creating systematic geographic mismatches between where disease risk is highest and where control capacity is greatest.

Although disease burden was consistently associated with insecticide deployment, deployment increased much more rapidly with increasing disease incidence in higher-HDI countries. This finding suggests that socioeconomic capacity strongly influences whether disease burden translates into large-scale implementation of vector-control interventions. Higher-HDI settings generally have greater capacity to procure insecticides, maintain surveillance systems, coordinate large-scale deployment, and implement insecticide resistance management.^29^ Furthermore, malaria control programs often benefit from substantial external financing through international initiatives, such as the Global Fund and bilateral donors, which can partially offset resource limitations while simultaneously introducing programmatic priorities that influence deployment patterns.^4^ Together, these findings indicate that disease burden alone does not determine deployment intensity, potentially creating systematic mismatches between where interventions are most needed and where they can be implemented at scale. The positive interaction between HDI and disease burden suggests that socioeconomic capacity influences not only baseline deployment but also how strongly countries are able to scale vector-control interventions in response to increasing disease risk. Collectively, these analyses suggest that vector-control deployment is governed by interacting epidemiological, socioeconomic, environmental, and operational constraints rather than by disease burden alone.

The intervention-specific analyses demonstrate that the determinants of insecticide deployment differ substantially among intervention types. ITNs and RS were most strongly associated with *Anopheles*-borne disease incidence, consistent with their long-standing roles in malaria control,^6^ whereas SS and LA exhibited stronger associations with *Aedes*-borne disease incidence and with one another, reflecting their complementary roles in arboviral vector control.^6^ The strong association between SS and LA suggests that these interventions are often deployed under similar epidemiological conditions. Combinations of infrastructure-intensive interventions, such as SS, RS, and LA, also exhibited strong socioeconomic dependence on HDI and population density, emphasizing that operational requirements differ substantially among vector-control strategies.^4^ For example, the positive interaction between population density and LA use for SS deployment and the positive interaction between HDI and RS use for LA deployment suggests that densely populated and high HDI settings can fully capitalize on the efficiencies of large-scale spraying and larviciding campaigns only when sufficient infrastructure and operational capacity are available.

The hurdle models also demonstrate that the factors determining whether an intervention is deployed are not always the same as those determining how extensively it is deployed once adopted. This distinction suggests that intervention adoption and deployment intensity are governed by different epidemiological, logistical, and financial processes, highlighting the importance of considering both decisions when evaluating and planning vector-control programs. Together, these findings are consistent with the integrated vector management framework advocated by the WHO Global Vector Control Response.^5^ Rather than treating insecticide deployment as a single planning problem, effective vector-control programs should tailor the combination of ITNs, RS, SS, and LA to local epidemiological, socioeconomic, and environmental conditions. Understanding how the drivers of deployment differ among interventions can help optimize integrated vector-control strategies rather than simply increasing deployment of all interventions simultaneously.

The climatic associations further suggest that environmental conditions influence both the need for and feasibility of insecticide deployment. Temperature and precipitation affect mosquito abundance and transmission potential,^9^ but they also influence the persistence of insecticides and the logistical challenges associated with implementation.^30^ The contrasting climatic responses observed among intervention types (for example, increased ITN deployment but reduced RS and SS deployment under wetter conditions) suggest that environmental conditions shape not only vector ecology but also which interventions are most practical and feasible in different settings.

The high-resolution prediction framework represents another important contribution of this study. National averages frequently masked substantial heterogeneity in predicted insecticide deployment among neighboring administrative units, demonstrating that coarse spatial summaries may overlook important local variation. The resulting ADM2 predictions provide a practical framework for identifying subnational regions where predicted deployment remains low relative to disease burden, providing predictions of locations where enhanced targeting by national vector-control programs, international donors, and sub-national control programs (e.g., states, municipalities) would be beneficial.

These predictions may help identify where intervention portfolios should differ among regions rather than assuming uniform deployment strategies within countries. As insecticide resistance can vary across regional mosquito populations, fine-scale deployment estimates may provide a stronger basis for adaptive management than national averages alone.

## Limitations

Several limitations warrant consideration. First, insecticide-use data were based on self-reported national records and were incomplete for some countries and years, potentially introducing reporting inconsistencies or measurement error. Second, our analyses relied on national-level associations to generate fine-scale predictions. Although this approach enables high-resolution mapping, it assumes that relationships estimated at the national scale apply locally; unmeasured subnational determinants may therefore reduce predictive accuracy in some regions. Third, our ecological analyses identify associations rather than causal relationships, and national socioeconomic and climatic indicators cannot fully capture the operational and programmatic factors that determine insecticide deployment. Similarly, the interactions identified here should be interpreted as evidence of context dependence rather than direct causal mechanisms and provide hypotheses for future mechanistic and operational studies. Additionally, local vector control programs may be using other proxies for insecticide deployment, such as vector density rather than disease incidence. Fourth, although our dataset captures the principal insecticide-based interventions targeting malaria and dengue vectors, it does not include other vector management approaches, including environmental management, housing improvements, biological control, vaccination campaigns, and therapeutics. Furthermore, while mosquitoes, sandflies, and triatomines overlap substantially at national scales, their distributions may differ locally, and our downscaled predictions may therefore underestimate deployment in regions where Chagas disease or leishmaniases occur in the absence of substantial mosquito-borne disease. Finally, standard spray coverage quantifies deployment rather than insecticide exposure, implementation quality, or intervention effectiveness and therefore should be interpreted as an indicator of deployment intensity rather than public health impact.

## Conclusions

Looking forward, the temporal dynamics of insecticide deployment in response to mosquito-borne disease, urbanization, and climate change represent important opportunities for future research. The continued spread of *Ae. albopictus* and *An. stephensi* is reshaping vector-borne disease risk by exposing new regions and rapidly growing urban centers to vectors that were previously absent.^2,3^ At the same time, rising temperatures and lengthening transmission seasons are unlikely to simply increase insecticide use uniformly; rather, they are expected to redistribute where interventions are needed, shifting the relative importance of ITNs, RS, SS, and LA across landscapes. For example, the ongoing invasion of *An. stephensi* into rapidly growing cities may increasingly require malaria interventions in urban environments, blurring these traditional distinctions.^3^ Predictive spatial frameworks such as those developed here could therefore be integrated with projected climatic, demographic, and urbanization scenarios to anticipate future intervention needs, identify emerging hotspots of insecticide demand, guide proactive resource allocation, and support insecticide resistance management before mismatches between disease risk and control capacity become more pronounced.

Overall, the broad geographic coverage, consistency of observed patterns, and cross-validated predictive performance suggest that this framework provides robust insights into global insecticide deployment and offers a practical tool for supporting vector-control planning. By integrating global surveillance data with predictive modeling and high-resolution mapping, our analyses identify where interventions are most likely to be deployed and where important deployment gaps remain. These spatially explicit predictions can support more equitable allocation of vector-control resources, guide insecticide resistance stewardship,^5^ and help national programs and international partners prioritize intervention strategies according to local epidemiological and infrastructural conditions. As mosquito distributions continue to shift under urbanization and changing environmental conditions, predictive mapping frameworks such as those presented here will become increasingly valuable for strengthening and sustaining both global and local vector-control programs.

## Contributors

PMH and JRR accessed and verified the underlying analytical data. HB verified the insecticide-use source data, and JM verified the insecticide-treated net source data. All authors had full access to the data relevant to their contributions, provided feedback on and edited the manuscript, approved the final manuscript, and approved the submission.

## Data sharing

The analytical dataset (with anonymized countries) and code necessary to reproduce the analyses are publicly available at https://github.com/pmheffernan/vector-targeted-insecticide-use. The de-anonymized dataset is available upon reasonable request.

## Disclosure statement on AI usage

During the preparation of this work, Google Gemini 3.6 Flash was used to troubleshoot errors encountered in the coding process. All Gemini suggestions were manually verified and tested for accuracy. The authors take full responsibility for the content of the published article.

## Acknowledgements

JRR acknowledges support from National Science Foundation (DEB-2109293, ITE-2333795, BCS-2307944, CISE-2435758), Notre Dame Poverty Initiative, Berthiaume Institute for Precision Health, IL IN Sea Grant, and Frontier Research Foundation. We are grateful to John Milliner and the Net Mapping Project for extracting and providing the insecticide-treated net dataset.

