## Supplement for "Global determinants of vector-targeted insecticide use in public health: a modeling and mapping analysis"

**Affiliations:**
[a] University of Notre Dame, Department of Biological Science, Environmental Change Initiative, Eck Institute of Global Health, Notre Dame, Indiana 46556, USA
[b] Laboratory of Entomology, Wageningen University, Wageningen 6700AA, Netherlands
[c] Academy of Public Health Entomology, Udaipur 313002, India
[d] Cornell University, Department of Entomology, 129 Garden Ave, Ithaca, 14850, NY, USA

Contents:

Supplementary Methods
Table S1. Regression coefficients, chi-squared values, and p-values for overall insecticide use model
Table S2. Regression coefficients, odds-ratios, chi-squared values, and p-values for binomial models of insecticide use types
Table S3. Regression coefficients, chi-squared values, and p-values for Gaussian models of insecticide use types
Table S4. Lasso Regression Lambda Values
Table S5. K-Folds CV Train and Test Mean $R^2$
Table S6. Regression coefficients, chi-squared values, and p-values for overall insecticide use model with vapor pressure deficit replacing temperature
Figure S1. Overall model responses not included in main text
Figure S2. ITN responses not included in main text
Figure S3. RS responses not included in main text
Figure S4. SS responses not included in main text
Figure S5. ADM0 map for overall standard spray coverage
Figure S6. ADM1 map for overall standard spray coverage
Figure S7. ADM2 map for insecticide use rate (standard spray coverage per person)
Figure S8. ADM1 map for insecticide use rate (standard spray coverage per person)
Figure S9. ADM0 map for insecticide use rate (standard spray coverage per person)
References

**Supplementary Methods**

*Insecticide use dataset*

Vector-targeting insecticide-use data were obtained from the global vector-control surveys[1,2] which report annual, national-level insecticide deployment by intervention type from 1990 through 2019. The datasets include insecticide-treated nets (ITNs), residual spraying (RS), spatial spraying (SS), and larviciding (LA; including both chemical and bacterial insecticides), with insecticide use reported as standard spray coverage (SSC; $km^2$ covered by active ingredient) to facilitate comparisons among intervention types. Observations from years in which insecticide use was not reported, or from countries that did not participate in the surveys, were excluded from the analyses.

Although these datasets include insecticide use targeting vectors of malaria, dengue, leishmaniases, and Chagas disease (among others), most reported insecticide use targeted malaria and dengue vectors. We therefore used *Anopheles*- and *Aedes*- borne disease incidences as proxies for the principal mosquito-borne diseases driving insecticide deployment. This approach is also justified because, at the national scale analyzed here, the geographic distributions of mosquitoes, sandflies, and triatomes often overlap, and insecticide applications targeting sandflies or triatomes are also likely to impose pressure on local mosquito populations.[3] Furthermore, high-resolution spatial incidence datasets required for our downscaling analyses are available for malaria and dengue but not for leishmaniases or Chagas disease. Consequently, *Anopheles*- and *Aedes*- borne disease incidence provided the most appropriate epidemiological predictors for both the statistical analyses and the spatial prediction framework.

The published insecticide-use datasets included ITN kits but excluded factory-treated ITNs. Following the methods of van den Berg et al. (2021a), annual country-level factory-treated ITN deliveries were obtained from the Net Mapping Project (J. Milliner, personal communication),[4] and corresponding SSC values were estimated using insecticide active ingredient and net dimensions before being added to the ITN and overall insecticide use datasets.

*Predictor variables*

For each country-year observation, predictor values corresponding to the year of reported insecticide use were extracted. Malaria incidence was used as a proxy for *Anopheles*-borne disease burden and dengue incidence as a proxy for *Aedes*-borne disease burden because these diseases dominate global insecticide-based control programs targeting their respective mosquito vectors. Disease incidence was defined as new human cases per 100,000 people and obtained from the Global Burden of Disease (GBD) database on October 12, 2025.[5]

Human Development Index (HDI) values were downloaded from the United Nations Development Programme[6] on May 08, 2025. Human population totals were obtained through World Bank[7] on October 12th, 2025 and divided by the land area reported in the Central Intelligence Agency's World Factbook[8] to obtain human population density (people per $km^2$).

Temperature (°C) and precipitation (m) were derived from the ERA5-Land monthly aggregated dataset from the European Centre for Medium-Range Weather Forecasts (ECMWF)[9] using Google Earth Engine.[10] Although mosquito distributions are influenced by additional climatic variables such as temperature extremes and seasonality, we focused on mean monthly temperature and precipitation because these variables are widely recognized as the primary environmental drivers of *Anopheles*- and *Aedes*-borne disease transmission.[11–15] They also influence insecticide deployment and effectiveness independently of vector population dynamics.[16–20] Because disease incidence was explicitly included in our models, insecticide-use observations were limited to countries implementing vector control, and analyses were conducted at the national scale, we prioritized model parsimony by restricting climatic predictors to these two variables. Due to increased evidence for the importance of humidity in both mosquito population dynamics[21,22] and insecticide persistence and efficacy,[23] we also tested the overall insecticide use model with mean monthly vapor pressure deficit (VPD; kPa) instead of temperature, which acts as a composite variable incorporating both temperature and relative humidity. VPD was also derived from the ERA5-Land monthly aggregated dataset[9] using the temperature and dew point temperature variables.

*Model development*

Overall insecticide use (i.e., combining ITN, RS, SS, and LA) was modeled using generalized linear mixed-effects models (GLMMs) implemented in the *glmmTMB*[24,25] package in *R*.[26] Country was included as a random intercept to account for repeated annual observations within countries, and national population (log-transformed) was included as an offset. Candidate fixed effects included *Anopheles*-borne disease incidence, *Aedes*-borne disease incidence, HDI, population density, mean monthly temperature, mean monthly precipitation, and all two- and three-way interactions among predictors.

*Anopheles*-borne disease incidence, *Aedes*-borne disease incidence, population density, precipitation, VPD, and insecticide use were log-transformed using *ln*(x+1), and the response variable, insecticide use, was transformed as *ln*(x+0.001). Predictors were subsequently scaled and centered to a mean of 0 and a standard deviation of 1. Temperature and precipitation (as well as VPD in the VPD model) were initially evaluated as second-order polynomial terms to allow for potential non-linearities. These terms did not improve model fit based on Bayesian information criterion (BIC) and were therefore excluded.

We then performed variable selection on the global model using lasso regression with the *glmmLasso*[27] package. To select the optimal *lambda* parameter for the lasso, we used five-folds cross-validation on a wide range of possible lambda values and selected the lambda with the lowest cross-validation deviance (Table S4). Predictors whose coefficients were shrunk to zero were removed. We fit the final model in *glmmTMB,* and model assumptions and quality of fit were assessed using *DHARMa.*[28] To assess model performance, we performed two types of repeated k-folds cross-validation (50 repeats) on the final model. First, to assess the model performance on trends within the dataset (the ability to downscale within known countries), we performed a cross-validation in which the folds (k=5) were split evenly among the countries. Second, to assess the model's predictive performance on unseen data (the ability to predict usage into regions outside the scope of the original dataset), we performed a cross-validation in which each fold (k=10) was split so that 90% of the countries only appeared in the training dataset and the remaining 10% of the countries only appeared in the testing dataset. Cross-validation results are presented in Table S5. Significance of each main effect and interaction was calculated using a type III ANOVA in the *car* package,[29] and marginal prediction plots for significant terms from the most parsimonious model were generated using *sjPlot*[30] and *ggplot.*[31]

Variable selection was performed using lasso (L1 penalized) regression implemented in the *glmmLasso* package.[27] The tuning parameter (*lambda*) was selected using five-fold cross-validation on a wide range of possible lambda values (tuning parameters reported in Table S4), and predictors whose coefficients were shrunk to zero were excluded from the final model. The reduced model was subsequently refitted in *glmmTMB,* model assumptions were evaluated using *DHARMa,*[28] and statistical significance was assessed using Type III ANOVA implemented in the *car* package.[29] The same overall process was followed for the model containing VPD instead of temperature. Marginal prediction plots were generated using *sjPlot*[30] and *ggplot.*[31]

*Hurdle models*

Separate hurdle models[32] were developed for each intervention type (ITN, RS, SS, and LA) because many countries reported zero use for individual interventions, whereas the combined insecticide-use dataset contained no structural zeroes. Each hurdle model consisted of a binomial component describing the probability of deployment and a Gaussian component describing the magnitude of deployment among countries reporting use.

In addition to the predictors included in the overall model, each hurdle model incorporated the use of the remaining three intervention types (SSC per person) as explanatory variables. Because of increased model complexity, only two-way interactions were evaluated. For each component of each hurdle model, variable selection, model reduction, and diagnostic procedures followed those used for the overall model. When lasso retained an interaction but eliminated its associated main effect, the corresponding main effect was retained in the final model according to the hierarchy principle.

For the LA hurdle model, *Aedes*-borne disease incidence was reintroduced after variable selection because of strong biological justification. Model fit was reassessed using BIC, after which this predictor was included because it did not decrease model performance.

*Cross-validation*

Predictive performance of the overall model was evaluated using two repeated cross-validation strategies (50 repetitions each). The first assessed interpolation within sampled countries by randomly partitioning observations among countries into five folds. The second assessed extrapolation to previously unseen countries by assigning 90% of countries to the training dataset and the remaining 10% to the testing dataset.

For hurdle models, predictive performance was evaluated using the same repeated cross-validation framework. Coefficients of determination ($R^2$) were calculated for Gaussian components, whereas area under the receiver operating characteristic curve (AUC) was calculated for binomial components. Model tuning parameters and cross-validation statistics are presented in Tables S4 and S5.

*Spatial prediction and mapping*

Whereas the use models are conducted at the country level, generating downscaled maps required equivalent inputs at finer spatial resolution. All spatial calculations were conducted using Google Earth Engine.[10] *Anopheles*-borne disease incidence was defined as the combined mean *Plasmodium falciparum* and *Plasmodium vivax* incidence per 100,000 people for each administrative region in 2024, obtained from the Malaria Atlas Project.[33] This approach does not capture non-falciparum or non-vivax malaria (e.g., *P. ovale*), which may contribute to insecticide deployment in some regions. *Aedes*-borne disease incidence was approximated using dengue incidence, as control efforts targeting other *Aedes*-borne diseases (e.g., Zika, chikungunya) typically overlap with dengue programs.[34] *Aedes*-borne disease incidence was estimated by combining dengue force of infection (FOI)[35] with LandScan population data.[36] Specifically, the total dengue incidence was calculated for each pixel by multiplying the dengue FOI layer by the population layer, and then these values were aggregated to the relevant administrative level. The total incidence was then divided by the population to obtain incidence rates (dengue cases per 100,000 people). Downscaled HDI values were obtained from Kummu et al's (2018),[37] human population density and the population offset were derived from LandScan,[36] and temperature and precipitation were calculated from ECMWF ERA5-Land Monthly Aggregated dataset averaged over 2000 to 2024.[9]

The overall insecticide use model was used to generate global prediction maps at the ADM0, ADM1, and ADM2 levels.[38] Using the *ggeffects* package[39] in *R,*[26] predicted values were generated from the overall model by coupling fitted model coefficients with the predictor values for each administrative unit. Administrative regions lacking one or more predictor variables were excluded. Predicted insecticide use was capped at the maximum SSC observed in the original insecticide-use dataset. To ensure predictions were constrained to epidemiologically relevant regions for mosquito-borne disease, predictions were only generated for countries with reported malaria or dengue incidence from 2014 to 2024 (the last ten years of the Global Burden of Disease dataset[5]). Furthermore, within those selected countries, predictions were only generated for regions that had a non-zero value for either *Anopheles*-borne disease incidence or *Aedes*-borne disease incidence. This reduced the risk of under-prediction in regions with non-*Aedes* or *Anopheles* vectors, such as sandflies and triatomes. The resulting ADM2 map displaying total SSC per administrative unit is presented in the main text, whereas the ADM0 and ADM1 maps as well as the maps displaying the SSC per person in each administrative unit are presented in the *Supplementary Materials*.

**Table S1 – Regression coefficients for overall insecticide use model**

| Predictors (scaled) | Overall Usage | | |
|---|---|---|---|
| | ***Coefficient (SE)*** | $\chi^2$ | ***p*** |
| Temperature | 0·346 (0·281) | 1·520 | 0·22 |
| Precipitation | -0·096 (0·198) | 0·233 | 0·63 |
| HDI | 0·947 (0·210) | 20·329 | < 0·0001 |
| Population | 0·121 (0·235) | 0·264 | 0·61 |
| AnBD | 0·904 (0·189) | 22·793 | < 0·0001 |
| AeBD | -0·001 (0·170) | 0 | 1 |
| Temperature*Precipitation | -0·409 (0·219) | 3·488 | 0·062 |
| Temperature*HDI | 0·412 (0·233) | 3·133 | 0·077 |
| Temperature*Population | -0·069 (0·276) | 0·063 | 0·8 |
| Temperature*AnBD | 0·371 (0·188) | 3·882 | 0·049 |
| Temperature*AeBD | -0·166 (0·201) | 0·686 | 0·41 |
| Precipitation*HDI | -0·270 (0·197) | 1·874 | 0·17 |
| Precipitation*Population | -0·010 (0·182) | 0·003 | 0·95 |
| Precipitation*AnBD | -0·374 (0·203) | 3·409 | 0·065 |
| Precipitation*AeBD | x | x | x |
| HDI*Population | 0·065 (0·176) | 0·135 | 0·71 |
| HDI*AnBD | 0·540 (0·139) | 15·072 | 0·0001 |
| HDI*AeBD | -0·003 (0·194) | 0 | 1 |
| Population*AnBD | 0·633 (0·200) | 10·033 | 0·0015 |
| Population*AeBD | -0·047 (0·161) | 0·086 | 0·77 |
| AnBD*AeBD | -0·427 (0·189) | 5·142 | 0·023 |
| Temperature*Precipitation*HDI | -0·212 (0·222) | 0·909 | 0·34 |
| Temperature*Precipitation*Population | -0·465 (0·222) | 4·368 | 0·04 |
| Temperature*Precipitation*AnBD | -0·143 (0·209) | 0·466 | 0·49 |
| Temperature*Precipitation*AeBD | -0·014 (0·136) | 0·010 | 0·92 |
| Temperature*HDI*Population | -0·508 (0·219) | 5·396 | 0·02 |
| Temperature*HDI*AnBD | -0·157 (0·129) | 1·491 | 0·22 |
| Temperature*HDI*AeBD | 0·201 (0·171) | 1·387 | 0·24 |
| Temeprature*Population*AnBD | -0·685 (0·268) | 6·546 | 0·01 |
| Temeprature*Population*AeBD | 0·033 (0·178) | 0·035 | 0·85 |
| Temeprature*AnBD*AeBD | x | x | x |
| Precipitation*HDI*Population | 0·290 (0·186) | 2·428 | 0·12 |
| Precipitation*HDI*AnBD | -0·080 (0·145) | 0·309 | 0·58 |
| Precipitation*HDI*AeBD | -0·081 (0·175) | 0·213 | 0·64 |
| Precipitation*Population*AnBD | 0·455 (0·227) | 4·003 | 0·045 |
| Precipitation*Population*AeBD | 0·214 (0·138) | 2·403 | 0·12 |
| Precipitation*AnBD*AeBD | 0·209 (0·152) | 1·891 | 0·169 |
| HDI*Population*AnBD | -0·067 (0·118) | 0·316 | 0·57 |
| HDI*Population*AeBD | -0·148 (0·171) | 0·748 | 0·39 |
| HDI*AnBD*AeBD | x | x | x |
| Population*AnBD*AeBD | -0·157 (0·129) | 0·152 | 0·7 |

*Note: "x" indicates a predictor variable that was selected out through lasso analysis. Temperature, precipitation, AnBD, AeBD, and population were each ln(x + 1) transformed· The response variable was ln(x + 0·001) transformed. Units: Temperature (degrees C), precipitation (meters), population (population density of people per sq-km), AeBD (Aedes-borne disease incidence [new cases per mid-year population per 100,000 people]), AnBD (Anopheles-borne disease incidence [new cases per mid-year population per 100,000 people]), and HDI (human development index). DF: The degrees of freedom for every variable and interaction is 1.*

**Table S2 – Regression coefficients for binomial models of insecticide-treated nets, residual spraying, space spraying, and larviciding**

| | ITN Use Likelihood | | | | RS Use Likelihood | | | | SS Use Likelihood | | | | LA Use Likelihood | | | |
|---|---|---|---|---|---|---|---|---|---|---|---|---|---|---|---|---|
| ***Predictors (scaled)*** | ***Coefficient (SE)*** | ***Odds Ratio*** | $\chi^2$ | ***p*** | ***Coefficient (SE)*** | ***Odds Ratio*** | $\chi^2$ | ***p*** | ***Coefficient (SE)*** | ***Odds Ratio*** | $\chi^2$ | ***p*** | ***Coefficient (SE)*** | ***Odds Ratio*** | $\chi^2$ | ***p*** |
| Temperature | 1·408 (0·655) | 4·088 | 4·619 | 0·032 | -1·288 (0·577) | 0·276 | 4·972 | 0·026 | 0·250 (0·620) | 1·284 | 0·163 | 0·67 | 0·575 (0·476) | 1·777 | 1·459 | 0·23 |
| Precipitation | 0·256 (0·376) | 1·292 | 0·463 | 0·5 | -0·357 (0·402) | 0·700 | 0·789 | 0·37 | 0·869 (0·421) | 2·385 | 4·267 | 0·039 | x | x | x | x |
| HDI | 2·040 (0·349) | 7·691 | 34·217 | < 0·0001 | -0·336 (0·424) | 0·715 | 0·628 | 0·43 | 0·500 (0·460) | 1·649 | 1·182 | 0·28 | 1·847 (0·357) | 6·341 | 26·738 | < 0·0001 |
| Population | 1·421 (0·589) | 4·141 | 5·818 | 0·016 | -0·423 (0·554) | 0·655 | 0·583 | 0·45 | 0·092 (0·583) | 1·096 | 0·025 | 0·87 | x | x | x | x |
| AnBD | 0·196 (0·387) | 1·217 | 0·256 | 0·61 | 1·157 (0·452) | 3·180 | 6·568 | 0·01 | -2·749 (0·585) | 0·064 | 22·086 | < 0·0001 | -1·319 (0·363) | 0·267 | 13·193 | 0·00028 |
| AeBD | 1·296 (0·306) | 3·655 | 17·959 | < 0·0001 | -0·449 (0·315) | 0·638 | 2·021 | 0·16 | 4·520 (0·550) | 91·836 | 67·415 | < 0·0001 | x | x | x | x |
| ITN | | | | | 0·301 (0·676) | 1·351 | 0·152 | 0·7 | 0·419 (0·801) | 1·520 | 0·273 | 0·6 | 0·333 (0·268) | 1·395 | 1·544 | 0·21 |
| RS | -0·086 (0·257) | 0·918 | 0·113 | 0·74 | | | | | -1·901 (1·013) | 0·149 | 3·525 | 0·06 | x | x | x | x |
| SS | 0·227 (0·696) | 1·255 | 0·106 | 0·74 | 0·380 (0·708) | 1·462 | 0·003 | 0·96 | | | | | 5·766 (1·654) | 319·258 | 12·158 | 0·00049 |
| LA | 0·010 (0·338) | 1·010 | 0·001 | 0·98 | -0·018 (0·948) | 0·982 | 0 | 0·98 | 3·779 (1·056) | 43·772 | 12·801 | 0·00035 | | | | |
| Temperature*Precipitation | x | x | x | x | x | x | x | x | x | x | x | x | x | x | x | x |
| Temperature*HDI | 1·143 (0·390) | 3·136 | 8·599 | 0·0034 | x | x | x | x | 0·166 (0·495) | 1·181 | 0·112 | 0·74 | x | x | x | x |
| Temperature*Population | x | x | x | x | x | x | x | x | -2·559 (0·632) | 0·077 | 16·380 | < 0·0001 | x | x | x | x |
| Temperature*AnBD | 1·925 (0·471) | 6·855 | 16·677 | 0·0034 | x | x | x | x | -1·727 (0·606) | 0·178 | 8·122 | 0·0044 | x | x | x | x |
| Temperature*AeBD | -0·674 (0·419) | 0·510 | 2·592 | 0·11 | x | x | x | x | x | x | x | x | x | x | x | x |
| Temperature*ITN | | | | | x | x | x | x | 1·175 (0·785) | 3·238 | 2·243 | 0·13 | x | x | x | x |
| Temperature*RS | -0·813 (0·316) | 0·444 | 6·603 | 0·01 | | | | | 0·688 (1·084) | 1·990 | 0·402 | 0·53 | x | x | x | x |
| Temperature*SS | -1·608 (0·892) | 0·200 | 3·255 | 0·07 | x | x | x | x | | | | | -1·755 (1·997) | 0·173 | 0·772 | 0·38 |
| Temperature*LA | x | x | x | x | 0·371 (0·602) | 1·449 | 0·380 | 0·54 | x | x | x | x | | | | |
| Precipitation*HDI | 0·786 (0·253) | 2·195 | 9·674 | 0·0019 | -2·201 (0·415) | 0·111 | 28·084 | < 0·0001 | -0·623 (0·355) | 0·536 | 3·080 | 0·079 | x | x | x | x |
| Precipitation*Population | x | x | x | x | x | x | x | x | x | x | x | x | x | x | x | x |
| Precipitation*AnBD | x | x | x | x | -2·165 (0·447) | 0·115 | 23·514 | < 0·0001 | -0·149 (0·415) | 0·862 | 0·129 | 0·72 | x | x | x | x |
| Precipitation*AeBD | -0·619 (0·204) | 0·538 | 9·209 | 0·0024 | x | x | x | x | -1·169 (0·329) | 0·311 | 12·590 | 0·00039 | x | x | x | x |
| Precipitation*ITN | | | | | -0·419 (0·186) | 0·658 | 5·078 | 0·024 | x | x | x | x | x | x | x | x |
| Precipitation*RS | x | x | x | x | | | | | 1·382 (0·486) | 3·983 | 8·083 | 0·0045 | x | x | x | x |
| Precipitation*SS | x | x | x | x | x | x | x | x | | | | | x | x | x | x |
| Precipitation*LA | x | x | x | x | x | x | x | x | x | x | x | x | | | | |
| HDI*Population | x | x | x | x | -0·149 (0·265) | 0·862 | 0·314 | 0·58 | -0·081 (0·289) | 0·922 | 0·078 | 0·78 | x | x | x | x |
| HDI*AnBD | 0·357 (0·238) | 1·429 | 2·248 | 0·13 | 3·047 (0·399) | 21·052 | 58·197 | < 0·0001 | -1·497 (0·373) | 0·224 | 16·118 | < 0·0001 | -1·551 (0·259) | 0·212 | 35·905 | < 0·0001 |
| HDI*AeBD | x | x | x | x | x | x | x | x | x | x | x | x | 0·425 (0·242) | 1·530 | 3·101 | 0·078 |
| HDI*ITN | | | | | 0·837 (0·498) | 2·309 | 2·820 | 0·093 | 0·069 (0·408) | 1·071 | 0·029 | 0·86 | 0·566 (0·283) | 1·761 | 4·006 | 0·045 |
| HDI*RS | x | x | x | x | | | | | -1·335 (1·130) | 0·263 | 1·396 | 0·24 | 2·304 (0·648) | 10·014 | 12·636 | 0·00038 |
| HDI*SS | x | x | x | x | -0·564 (0·774) | 0·569 | 0·530 | 0·47 | | | | | x | x | x | x |
| HDI*LA | x | x | x | x | x | x | x | x | 2·613 (0·592) | 13·640 | 19·455 | < 0·0001 | | | | |

**Table S2 (Continued)**

| Predictors (scaled) | ITN Use Likelihood | | | | RS Use Likelihood | | | | SS Use Likelihood | | | | LA Use Likelihood | | | |
|---|---|---|---|---|---|---|---|---|---|---|---|---|---|---|---|---|
| | *Coefficient (SE)* | *Odds Ratio* | $\chi^2$ | *p* | *Coefficient (SE)* | *Odds Ratio* | $\chi^2$ | *p* | *Coefficient (SE)* | *Odds Ratio* | $\chi^2$ | *p* | *Coefficient (SE)* | *Odds Ratio* | $\chi^2$ | *p* |
| Population*AnBD | 0·649 (0·310) | 1·914 | 4·392 | 0·036 | x | x | x | x | x | x | x | x | x | x | x | x |
| Population*AeBD | x | x | x | x | x | x | x | x | 1·107 (0·391) | 3·025 | 8·001 | 0·0047 | x | x | x | x |
| Population*ITN | | | | | x | x | x | x | 0·691 (0·362) | 1·996 | 3·651 | 0·056 | x | x | x | x |
| Population*RS | 0·225 (0·166) | 1·252 | 1·826 | 0·18 | | | | | -3·558 (1·016) | 0·028 | 12·257 | 0·00046 | x | x | x | x |
| Population*SS | -1·562 (0·636) | 0·210 | 6·033 | 0·014 | x | x | x | x | | | | | x | x | x | x |
| Population*LA | -0·710 (0·319) | 0·492 | 4·946 | 0·026 | x | x | x | x | 0·286 (0·254) | 1·331 | 1·269 | 0·26 | | | | |
| AnBD*AeBD | x | x | x | x | x | x | x | x | 2·445 (0·405) | 11·531 | 36·373 | < 0·0001 | x | x | x | x |
| AnBD*ITN | | | | | x | x | x | x | x | x | x | x | x | x | x | x |
| AnBD*RS | -0·266 (0·227) | 0·766 | 1·381 | 0·24 | | | | | -5·400 (1·587) | 0·005 | 11·581 | 0·00067 | x | x | x | x |
| AnBD*SS | -0·101 (0·300) | 0·904 | 0·113 | 0·74 | -0·095 (0·734) | 0·909 | 0·017 | 0·90 | | | | | x | x | x | x |
| AnBD*LA | x | x | x | x | x | x | x | x | x | x | x | x | | | | |
| AeBD*ITN | | | | | x | x | x | x | x | x | x | x | -0·261 (0·196) | 0·770 | 1·782 | 0·18 |
| AeBD*RS | -0·159 (0·167) | 0·853 | 0·906 | 0·34 | | | | | 0·055 (0·755) | 1·057 | 0·0052 | 0·94 | x | x | x | x |
| AeBD*SS | x | x | x | x | 0·291 (0·431) | 1·338 | 0·456 | 0·5 | | | | | x | x | x | x |
| AeBD*LA | 0·183 (0·288) | 1·201 | 0·404 | 0·53 | x | x | x | x | 5·275 (1·160) | 195·39 | 20·677 | < 0·0001 | | | | |
| ITN*RS | | | | | | | | | 0·307 (0·212) | 1·359 | 2·093 | 0·15 | x | x | x | x |
| ITN*SS | | | | | 0·081 (1·090) | 1·084 | 0·006 | 0·94 | | | | | x | x | x | x |
| ITN*LA | | | | | -2·508 (3·354) | 0·081 | 0·559 | 0·45 | 7·100 (3·715) | 1211·967 | 3·653 | 0·056 | | | | |
| RS*SS | -0·328 (0·472) | 0·720 | 0·482 | 0·49 | | | | | | | | | x | x | x | x |
| RS*LA | x | x | x | x | | | | | -2·525 (0·565) | 0·08 | 19·941 | < 0·0001 | | | | |
| SS*LA | x | x | x | x | x | x | x | x | | | | | | | | |

*Note: A blank cell indicates a predictor variable that was not tested in the model. "x" indicates a predictor variable that was selected out through lasso analysis. Temperature, precipitation, AnBD, AeBD, and population were ln(x + 1) transformed. All predictors were scaled and centered.*
*Units: Temperature (degrees C), precipitation (meters), population (density; people per sq-km), AeBD (Aedes-borne disease incidence [new cases per mid-year population per 100,000 people]), AnBD (Anopheles-borne disease incidence [new cases per mid-year population per 100,000 people]), HDI (human development index), ITN (insecticide-treated nets, standard spray coverage [SSC] per sq-km), RS (residual spraying, SSC per sq-km), SS (space spraying, SSC per sq-km), and LA (larviciding, SSC per sq-km).*

*DF: The degrees of freedom for every variable and interation in all models is 1.*

**Table S3 – Regression coefficients for Gaussian models of insecticide-treated nets, residual spraying, space spraying, and larviciding**

| | ITN Use | | | RS Use | | | SS Use | | | LA Use | | |
|---|---|---|---|---|---|---|---|---|---|---|---|---|
| ***Predictors (scaled)*** | ***Coefficient (SE)*** | $\chi^2$ | $p$ | ***Coefficient (SE)*** | $\chi^2$ | $p$ | ***Coefficient (SE)*** | $\chi^2$ | $p$ | ***Coefficient (SE)*** | $\chi^2$ | $p$ |
| Temperature | 1·063 (0·354) | 9·037 | 0·0026 | x | x | x | x | x | x | x | x | x |
| Precipitation | 0·201 (0·262) | 0·590 | 0·44 | 0·009 (0·153) | 0·003 | 0·95 | 0·298 (0·161) | 3·429 | 0·064 | x | x | x |
| HDI | 1·547 (0·256) | 36·337 | < 0·0001 | -0·375 (0·146) | 6·540 | 0·011 | 0·170 (0·205) | 0·688 | 0·41 | -0·094 (0·166) | 0·321 | 0·57 |
| Population | 0·575 (0·339) | 2·871 | 0·09 | -0·022 (0·203) | 0·012 | 0·91 | -0·221 (0·224) | 0·972 | 0·32 | x | x | x |
| AnBD | 0·757 (0·300) | 6·362 | 0·01 | 0·327 (0·152) | 4·634 | 0·031 | -0·185 (0·222) | 0·694 | 0·4 | -0·486 (0·163) | 8·852 | 0·0029 |
| AeBD | -0·226 (0·200) | 1·276 | 0·26 | x | x | x | 0·150 (0·135) | 1·238 | 0·266 | 0·286 (0·108) | 6·985 | 0·0082 |
| ITN | | | | 0·059 (0·141) | 0·172 | 0·68 | x | x | x | x | x | x |
| RS | x | x | x | | | | x | x | x | 0·132 (0·075) | 3·149 | 0·076 |
| SS | -0·147 (0·318) | 0·212 | 0·65 | x | x | x | | | | 0·014 (0·038) | 0·131 | 0·72 |
| LA | -0·070 (0·191) | 0·134 | 0·71 | 0·083 (0·172) | 0·230 | 0·63 | 0·041 (0·083) | 0·246 | 0·62 | | | |
| Temperature*Precipitation | x | x | x | x | x | x | x | x | x | x | x | x |
| Temperature*HDI | x | x | x | x | x | x | x | x | x | x | x | x |
| Temperature*Population | x | x | x | x | x | x | x | x | x | x | x | x |
| Temperature*AnBD | x | x | x | x | x | x | x | x | x | x | x | x |
| Temperature*AeBD | x | x | x | x | x | x | x | x | x | x | x | x |
| Temperature*ITN | | | | x | x | x | x | x | x | x | x | x |
| Temperature*RS | -0·722 (0·354) | 4·156 | 0·04 | | | | x | x | x | x | x | x |
| Temperature*SS | x | x | x | x | x | x | | | | x | x | x |
| Temperature*LA | x | x | x | x | x | x | x | x | x | | | |
| Precipitation*HDI | x | x | x | -0·360 (0·106) | 11·520 | 0·00069 | x | x | x | x | x | x |
| Precipitation*Population | x | x | x | x | x | x | x | x | x | x | x | x |
| Precipitation*AnBD | x | x | x | x | x | x | x | x | x | x | x | x |
| Precipitation*AeBD | x | x | x | x | x | x | x | x | x | x | x | x |
| Precipitation*ITN | | | | x | x | x | x | x | x | x | x | x |
| Precipitation*RS | 0·301 (0·082) | 13·579 | 0·00022 | | | | x | x | x | x | x | x |
| Precipitation*SS | x | x | x | x | x | x | | | | x | x | x |
| Precipitation*LA | x | x | x | x | x | x | -0·260 (0·088) | 8·773 | 0·0031 | | | |
| HDI*Population | -0·184 (0·198) | 0·859 | 0·35 | x | x | x | x | x | x | x | x | x |
| HDI*AnBD | 1·004 (0·194) | 26·743 | < 0·0001 | 0·519 (0·109) | 22·615 | < 0·0001 | x | x | x | x | x | x |
| HDI*AeBD | 0·559 (0·160) | 12·167 | 0·00049 | x | x | x | 0·233 (0·160) | 2·116 | 0·15 | x | x | x |
| HDI*ITN | | | | x | x | x | x | x | x | x | x | x |
| HDI*RS | x | x | x | | | | x | x | x | 0·347 (0·129) | 7·214 | 0·0072 |
| HDI*SS | x | x | x | x | x | x | | | | x | x | x |
| HDI*LA | x | x | x | x | x | x | x | x | x | | | |

**Table S3 (Continued)**

| Predictors (scaled) | ITN Use | | | RS Use | | | SS Use | | | LA Use | | |
|---|---|---|---|---|---|---|---|---|---|---|---|---|
| | ***Coefficient (SE)*** | $\chi^2$ | *p* | ***Coefficient (SE)*** | $\chi^2$ | *p* | ***Coefficient (SE)*** | $\chi^2$ | *p* | ***Coefficient (SE)*** | $\chi^2$ | *p* |
| Population*AnBD | x | x | x | x | x | x | x | x | x | x | x | x |
| Population*AeBD | x | x | x | x | x | x | x | x | x | x | x | x |
| Population*ITN | | | | 0·200 (0·087) | 5·230 | 0·022 | x | x | x | x | x | x |
| Population*RS | x | x | x | | | | x | x | x | x | x | x |
| Population*SS | x | x | x | x | x | x | | | | x | x | x |
| Population*LA | x | x | x | x | x | x | 0·180 (0·072) | 6·223 | 0·013 | | | |
| AnBD*AeBD | x | x | x | x | x | x | -0·124 (0·153) | 0·658 | 0·42 | x | x | x |
| AnBD*ITN | | | | x | x | x | x | x | x | x | x | x |
| AnBD*RS | 0·204 (0·211) | 0·937 | 0·33 | | | | x | x | x | x | x | x |
| AnBD*SS | x | x | x | x | x | x | | | | -0·361 (0·087) | 17·359 | 0·0072 |
| AnBD*LA | x | x | x | -0·136 (0·038) | 12·955 | 0·00032 | x | x | x | | | |
| AeBD*ITN | | | | x | x | x | x | x | x | x | x | x |
| AeBD*RS | x | x | x | | | | x | x | x | x | x | x |
| AeBD*SS | x | x | x | x | x | x | | | | x | x | x |
| AeBD*LA | x | x | x | x | x | x | x | x | x | | | |
| ITN*RS | | | | | | | x | x | x | x | x | x |
| ITN*SS | | | | x | x | x | | | | x | x | x |
| ITN*LA | | | | -0·195 (0·697) | 0·078 | 0·78 | x | x | x | | | |
| RS*SS | -0·064 (0·669) | 0·009 | 0·92 | | | | | | | x | x | x |
| RS*LA | 0·857 (0·479) | 3·199 | 0·074 | | | | x | x | x | | | |
| SS*LA | x | x | x | x | x | x | | | | | | |

*Note: A blank cell indicates a predictor variable that was not tested in the model. "x" indicates a predictor variable that was selected out through lasso analysis. Temperature, precipitation, AnBD, AeBD, and population were ln(x + 1) transformed, each response was ln(x + 0·001) transformed. All predictors were scaled and centered.*

*Units: Temperature (degrees C), precipitation (meters), population (density; people per sq-km), AeBD (Aedes-borne disease incidence [new cases per mid-year population per 100,000 people]), AnBD (Anopheles-borne disease incidence [new cases per mid-year population per 100,000 people]), HDI (human development index), ITN (insecticide-treated nets, standard spray coverage [SSC] per sq-km), RS (residual spraying, SSC per sq-km), SS (space spraying, SSC per sq-km), and LA (larviciding, SSC per sq-km).*

*DF: The degrees of freedom for every variable and interation in all models is 1.*

**Table S4. Lasso Regression Lambda Values.**

| Model | Lambda |
|---|---|
| Overall | 7·166 |
| Overall (*VPD*) | 2·151 |
| ITN (Binom.) | 5·045 |
| ITN (Contin.) | 42·785 |
| RS (Binom.) | 7·520 |
| RS (Contin.) | 51·995 |
| SS (Binom.) | 5·539 |
| SS (Contin.) | 52·161 |
| LA (Binom.) | 10·579 |
| LA (Contin.) | 60·654 |

**Table S5. K-Folds CV Train and Test Mean $R^2$.** For the overall model, *O* represents the results for cross-validation performed on folds holding out 10% of countries, and *I* represents the results for cross-validation performed on folds split evenly among the countries. For the intervention-specific models, only the *I* cross validation was performed. *VPD model* refers to the overall annual insecticide use model testing vapor pressure deficit (VPD) instead of temperature.

| Model | Mean Train $R^2$/AUC | Mean Test $R^2$/AUC |
|---|---|---|
| Overall (O) | 0·706 (*conditional* $R^2$)<br>0·461 (*marginal* $R^2$) | 0·327 ($R^2$) |
| Overall (I) | 0·756 (*conditional* $R^2$)<br>0·570 (*marginal* $R^2$) | 0·522 ($R^2$) |
| Overall (O)<br>*VPD model* | 0·709 (*conditional* $R^2$)<br>0·464 (*marginal* $R^2$) | 0·351 ($R^2$) |
| Overall (I)<br>*VPD model* | 0·776 (*conditional* $R^2$)<br>0·562 (*marginal* $R^2$) | 0·534 ($R^2$) |
| ITN (Binom.) | 0·923 (AUC) | 0·818 (AUC) |
| ITN (Contin.) | 0·656 ($R^2$) | 0·299 ($R^2$) |
| RS (Binom.) | 0·994 (AUC) | 0·884 (AUC) |
| RS (Contin.) | 0·856 ($R^2$) | 0·665 ($R^2$) |
| SS (Binom.) | 0·990 (AUC) | 0·898 (AUC) |
| SS (Contin.) | 0·849 ($R^2$) | 0·626 ($R^2$) |
| LA (Binom.) | 0·979 (AUC) | 0·905 (AUC) |
| LA (Contin.) | 0·871 ($R^2$) | 0·646 ($R^2$) |

**Table S6 – Regression coefficients for overall VPD insecticide use model**

| | Overall Usage | | |
|---|---|---|---|
| ***Predictors (scaled)*** | ***Coefficient (SE)*** | $\chi^2$ | ***p*** |
| VPD | 0·777 (0·357) | 4·750 | 0·029 |
| Precipitation | 0·097 (0·351) | 0·076 | 0·78 |
| HDI | 0·919 (0·255) | 13·026 | 0·0003 |
| Population | -0·167 (0·262) | 0·407 | 0·52 |
| AnBD | 0·515 (0·247) | 4·343 | 0·037 |
| AeBD | 0·081 (0·201) | 0·160 | 0·69 |
| VPD*Precipitation | -0·141 (0·212) | 0·445 | 0·5 |
| VPD*HDI | 0·372 (0·370) | 1·016 | 0·31 |
| VPD*Population | -0·017 (0·363) | 0·002 | 0·96 |
| VPD*AnBD | 0·353 (0·365) | 0·935 | 0·33 |
| VPD*AeBD | -0·194 (0·264) | 0·537 | 0·46 |
| Precipitation*HDI | -0·395 (0·380) | 1·076 | 0·3 |
| Precipitation*Population | -0·322 (0·322) | 0·998 | 0·32 |
| Precipitation*AnBD | -0·460 (0·369) | 1·559 | 0·21 |
| Precipitation*AeBD | -0·343 (0·236) | 2·115 | 0·15 |
| HDI*Population | 0·104 (0·167) | 0·389 | 0·53 |
| HDI*AnBD | 0·679 (0·148) | 21·031 | < 0·0001 |
| HDI*AeBD | 0·292 (0·174) | 2·799 | 0·094 |
| Population*AnBD | 0·564 (0·182) | 9·596 | 0·002 |
| Population*AeBD | -0·040 (0·144) | 0·075 | 0·78 |
| AnBD*AeBD | -0·128 (0·180) | 0·505 | 0·48 |
| VPD*Precipitation*HDI | -0·387 (0·245) | 2·499 | 0·11 |
| VPD*Precipitation*Population | -0·248 (0·226) | 1·211 | 0·27 |
| VPD*Precipitation*AnBD | -0·725 (0·243) | 8·924 | 0·0028 |
| VPD*Precipitation*AeBD | -0·065 (0·162) | 0·160 | 0·69 |
| VPD*HDI*Population | 0·313 (0·265) | 1·396 | 0·24 |
| VPD*HDI*AnBD | -0·360 (0·187) | 3·708 | 0·054 |
| VPD*HDI*AeBD | 0·493 (0·266) | 3·432 | 0·064 |
| VPD*Population*AnBD | 0·004 (0·311) | 0 | 0·99 |
| VPD*Population*AeBD | -0·461 (0·214) | 4·648 | 0·031 |
| VPD*AnBD*AeBD | 0·667 (0·247) | 7·262 | 0·007 |
| Precipitation*HDI*Population | 0·604 (0·289) | 4·377 | 0·036 |
| Precipitation*HDI*AnBD | -0·617 (0·236) | 6·832 | 0·009 |
| Precipitation*HDI*AeBD | 0·116 (0·259) | 3·432 | 0·064 |
| Precipitation*Population*AnBD | 0·580 (0·346) | 2·811 | 0·094 |
| Precipitation*Population*AeBD | -0·250 (0·217) | 1·329 | 0·25 |
| Precipitation*AnBD*AeBD | 0·240 (0·202) | 1·411 | 0·23 |
| HDI*Population*AnBD | -0·264 (0·130) | 4·145 | 0·042 |
| HDI*Population*AeBD | -0·333 (0·174) | 3·650 | 0·056 |
| HDI*AnBD*AeBD | 0·236 (0·149) | 2·503 | 0·11 |
| Population*AnBD*AeBD | -0·194 (0·174) | 1·241 | 0·27 |

*Note: "x" indicates a predictor variable that was selected out through lasso analysis. Vapor pressure deficit (VPD), precipitation, AnBD, AeBD, and population were each ln(x + 1) transformed· The response variable was ln(x + 0·001) transformed. Units: Temperature (degrees C), precipitation (meters), population (population density of people per sq-km), AeBD (Aedes-borne disease incidence [new cases per mid-year population per 100,000 people]), AnBD (Anopheles-borne disease incidence [new cases per mid-year population per 100,000 people]), and HDI (human development index). DF: The degrees of freedom for every variable and interaction is 1.*

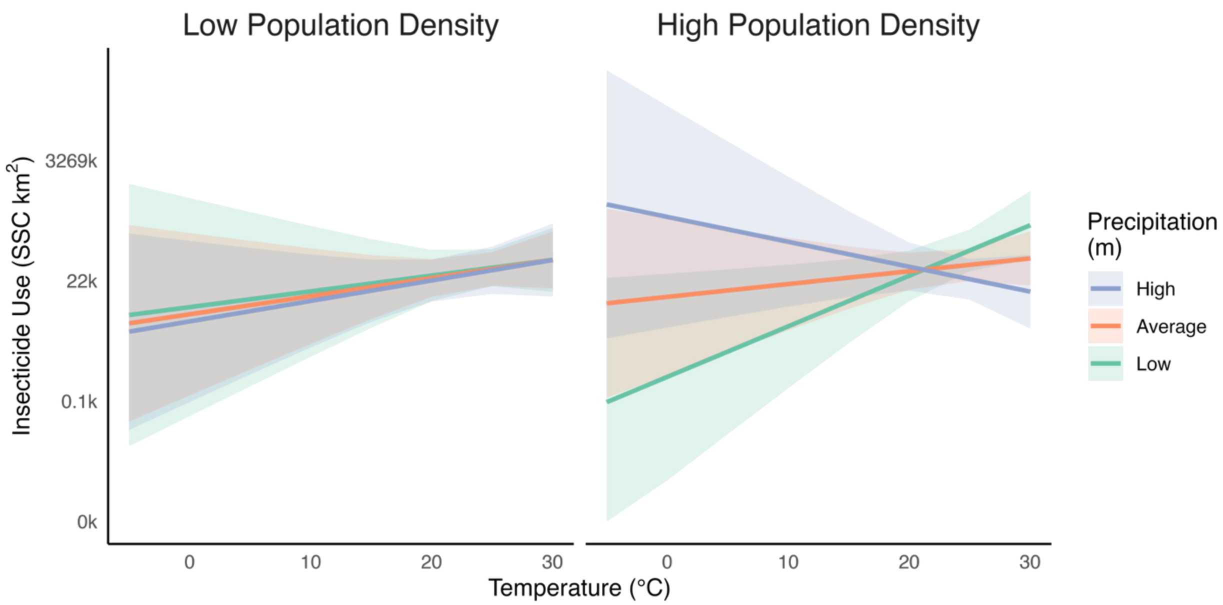


**Figure S1.** Marginal predictive plots displaying significant associations from the overall annual insecticide (SSC = standard spray coverage in $km^2$) use model not discussed in the main text, with temperature, precipitation, and population density as predictors. Insecticide use and precipitation are presented on a natural log scale. The best fit lines and 95% confidence bands are displayed. *High* and *Low* represent the +/- 1 standard deviation from the mean.

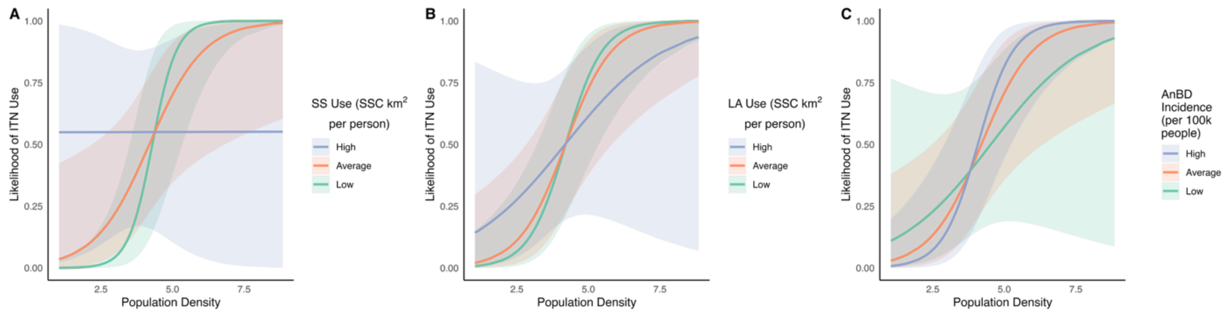


**Figure S2.** Marginal predictive plots displaying significant associations from the likelihood component of the annual insecticide-treated net (ITN; SSC = standard spray coverage in km$^2$) use hurdle model not discussed in the main text, with population density (people per km$^2$, **A-C**), space spraying (SS) use (SSC per person; **A**), larvicide (LA) use (SSC per person; **B**), and *Anopheles*-borne disease incidence (AnBD; new cases divided by mid-year population; **C**) as predictors. Population density, LA use, RS use, and AnBD incidence are presented on a natural log scale. The best fit lines and 95% confidence bands are displayed. In figures with interactions, *High* and *Low* represent the +/- 1 standard deviation from the mean.

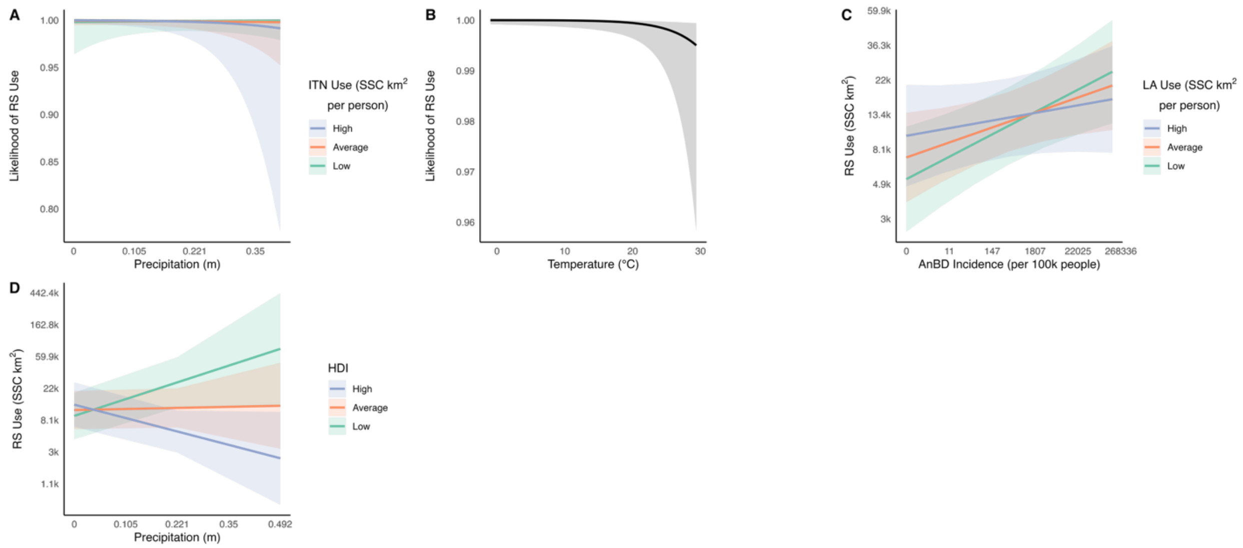


**Figure S3.** Marginal predictive plots displaying significant associations from the likelihood (**A-B**) and magnitude (**C-D**) components of the annual residual spraying (RS; SSC = standard spray coverage in km$^2$) use hurdle model not discussed in the main text, with precipitation (m; mean monthly; **A, D**), insecticide-treated net (ITN) use (SSC per person; **A**), temperature (mean monthly, °C; **B**), *Anopheles*-borne disease incidence (AnBD; new cases divided by mid-year population; **C**), larvicide (LA) use (SSC per person; **C**), and human development index (HDI; **D**) as predictors. RS Use, ITN use, LA use, AnBD incidence, and precipitation are presented on a natural log scale. The best fit lines and 95% confidence bands are displayed. In figures with interactions, *High* and *Low* represent the +/- 1 standard deviation from the mean.

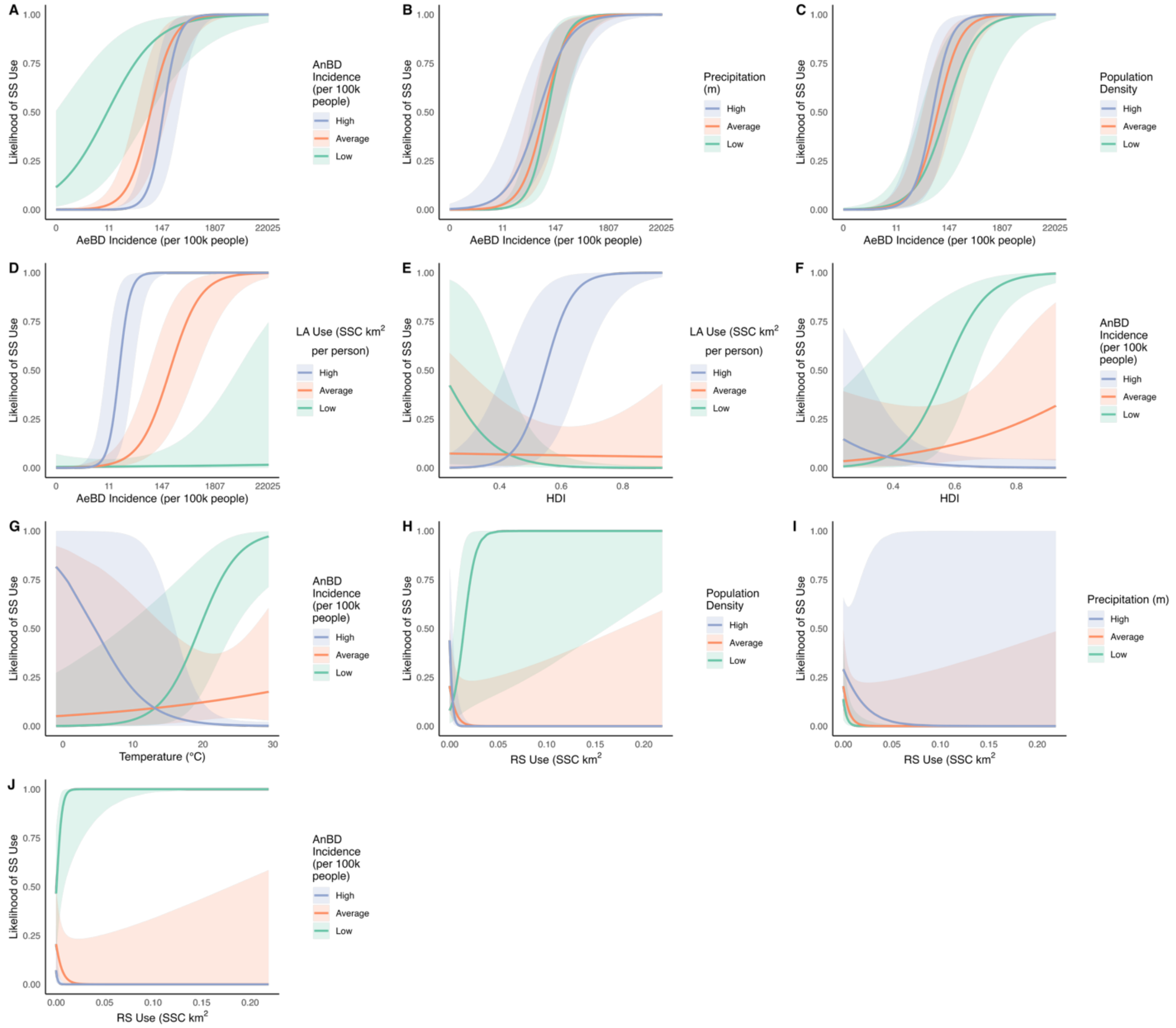


**Figure S4.** Marginal predictive plots displaying significant associations from the likelihood component of the annual space spraying (SS; SSC = standard spray coverage in km$^2$) use hurdle model not discussed in the main text, with *Aedes*-borne disease incidence (AeBD; new cases divided by mid-year population; **A-D**), *Anopheles*-borne disease incidence (AnBD; new cases divided by mid-year population; **A, F-G, J**), precipitation (mean monthly, m; **B, I**), population density (people per km$^2$; **C, H**), larvicide (LA) use (SSC per person; **D-E**), human development index (HDI; **E-F**), temperature (mean monthly, °C; **G**), and residual spraying (RS) use (SSC per person; **H-J**) as predictors. SS Use, LA Use, RS use, AeBD incidence, AnBD incidence, population density, and precipitation are presented on a natural log scale. The best fit lines and 95% confidence bands are displayed. In figures with interactions, High and Low represent the +/- 1 standard deviation from the mean.

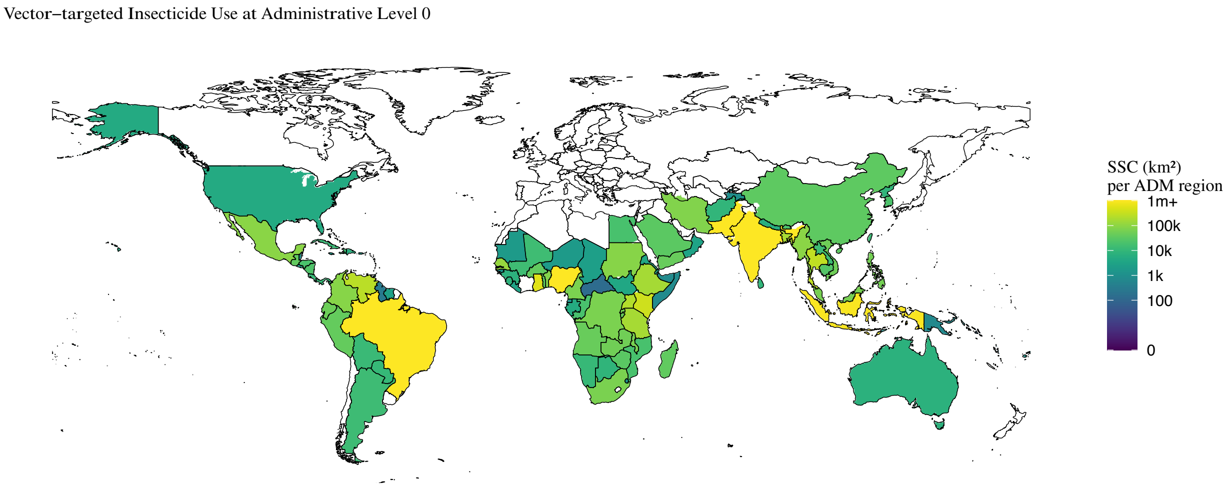


**Figure S5.** Predicted magnitude of total annual insecticide use (standard spray coverage [SSC] in $km^2$; including insecticide-treated nets, residual spraying, spatial spraying, and larvicide) for *Anopheles*- and *Aedes*-borne disease control at the administrative level 0 (ADM0).

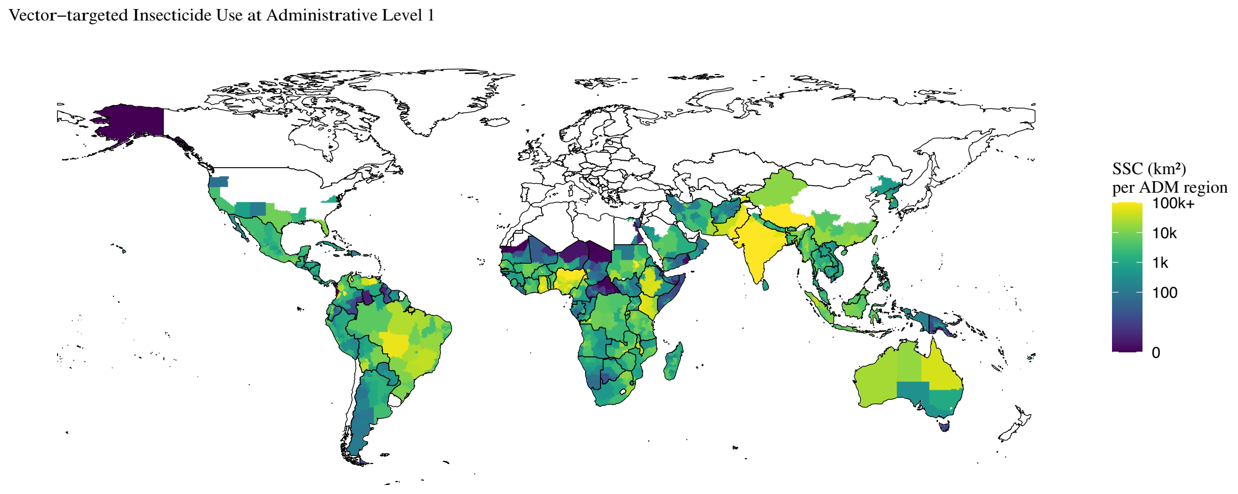


**Figure S6.** Predicted magnitude of total annual insecticide use (standard spray coverage [SSC] in km$^2$) including insecticide-treated nets, residual spraying, spatial spraying, and larvicide) for *Anopheles*- and *Aedes*-borne disease control at the administrative level 1 (ADM1).

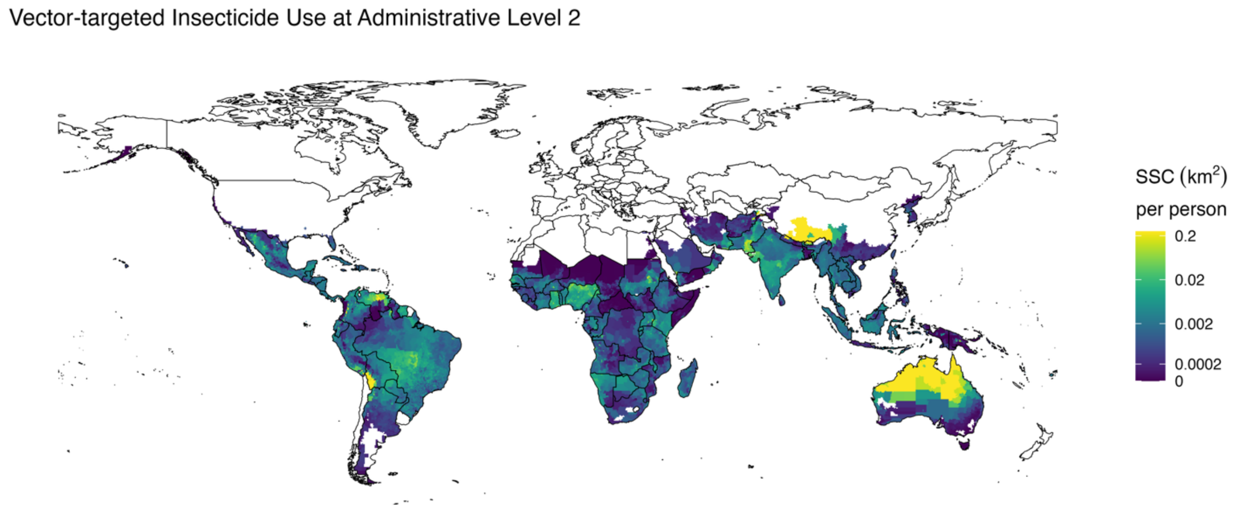


**Figure S7.** Predicted annual insecticide use rate (standard spray coverage in $km^2$ per person; including insecticide-treated nets, residual spraying, spatial spraying, and larvicide) for *Anopheles*- and *Aedes*-borne disease control at the administrative level 2 (ADM2).

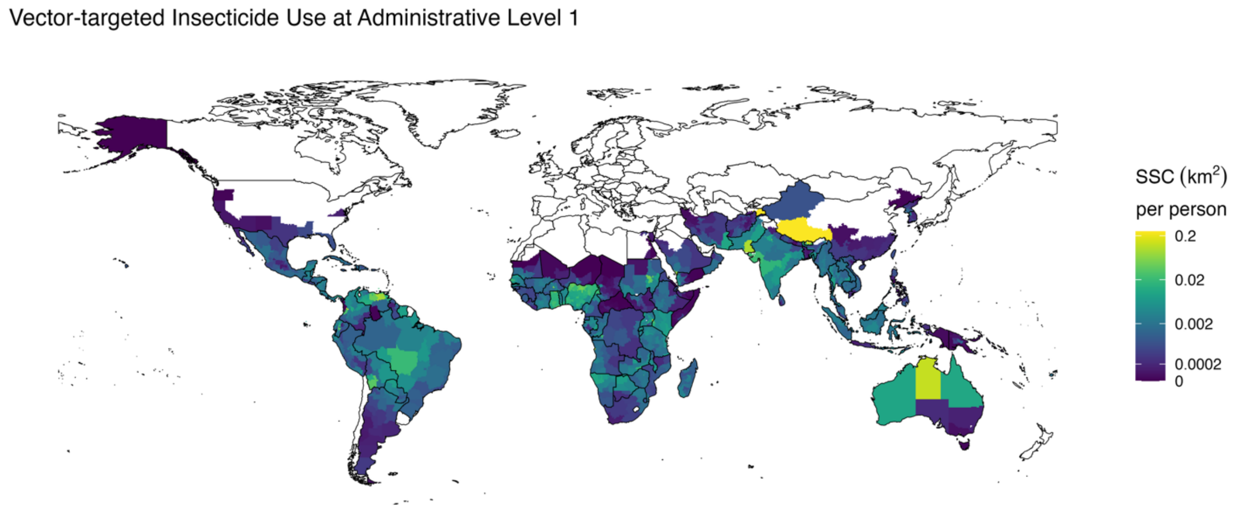


**Figure S8.** Predicted annual insecticide use rate (standard spray coverage in km$^2$ per person; including insecticide-treated nets, residual spraying, spatial spraying, and larviciding) for *Anopheles*- and *Aedes*-borne disease control at the administrative level 1 (ADM1).

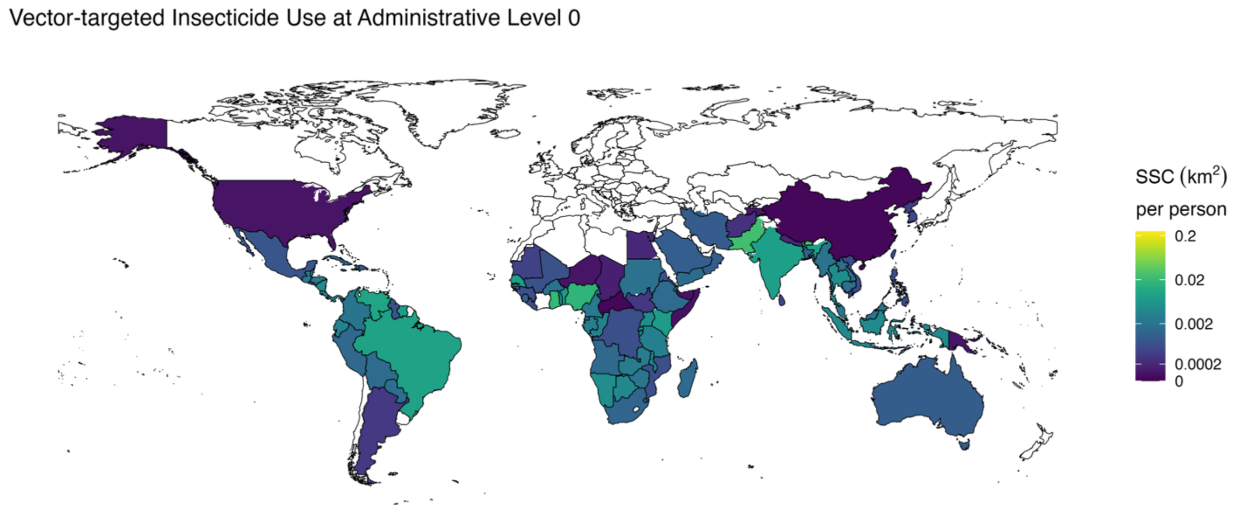


**Figure S9.** Predicted annual insecticide use rate (standard spray coverage in $km^2$ per person; including insecticide-treated nets, residual spraying, spatial spraying, and larviciding) for *Anopheles*- and *Aedes*-borne disease control at the administrative level 0 (ADM0).

5. Global Burden of Disease Collaborative Network. Global Burden of Disease Study 2023 (GBD 2023). *Seattle U S Inst Health Metr Eval IHME*. Published online 2025.
